# Validating methods for inferring co-occurring diseases: a flexible framework for simulating synthetic data

**DOI:** 10.64898/2026.01.13.26344018

**Authors:** Hannah Marchi, Sophie Schmiegel, Tamara Schamberger, Christiane Fuchs

## Abstract

The validation of methods is an integral part of statistical research, defining conditions under which methods can yield reliable results. When validation is carried out empirically, it requires a solid data basis that allows the control and management of relevant characteristics. In particular, depending on the context, the data must meet specific requirements regarding, e. g. sample size, dimensionality, completeness and underlying dependency structures. Real-world data often fails to meet these requirements, particularly in medical contexts where availability or the right to publish is additionally restricted through privacy regulations. For this reason, synthetic data is an effective alternative for method validation.

Generating synthetic data is particularly demanding if it is required to precisely mirror complex dependence structures while simultaneously controlling certain target characteristics. In our work, we address the medical context of simultaneously co-occurring diseases, where symptoms may overlap or conflict. We seek to generate synthetic data supporting the simulation-based validation of statistical methods which are able to predict holistic disease pictures based on patient information, including the accounting for comorbidities. We introduce a four-step framework in which we (I) generate patient covariates such as symptoms; (II) connect this patient information to predictors for the single or joint occurrence of diseases; (III) transform the predictors into disease probabilities or disease scores; (IV) convert the probabilities or scores into disease occurrence. Within each of these steps, we outline several alternatives that allow different forms of modeling the overall dependence structure.

We apply our framework to a case study informed by real-world data: to the context of pain-causing diseases which share certain similarities in their clinical presentations, and which can occur either individually or jointly. In this analysis, we employ five combinations of methodological alternatives within the data generation steps. This allows us to evaluate the data generation approaches with respect to their ability to achieve the defined target characteristics, and to demonstrate strengths and weaknesses as well as specific suitability.

The proposed simulation framework is broadly applicable beyond the specific use case and the medical context. The approaches are designed to be accessible and adjustable through varying input settings, enabling users to tailor the data generation to their specific needs. This way, our work provides researchers with a flexible framework for generating synthetic validation data that aligns with the methodological requirements of their studies.

## 1 Introduction

A relevant part of statistics is inferential statistics, where conclusions are drawn from a data sample about an underlying population. An important subarea within this field is the discovery and learning of relationships between observed or unobserved random variables. To study such relationships, a wide range of statistical methods exists, where the suitability of each method depends on the assumptions about the underlying ground truth as well as on the availability and characteristics of the data. It frequently occurs that problems of interest are structured in such a way that existing inference methods cannot be applied directly, which leads to the development of new approaches. For both newly developed and already established methods, method validation is a crucial step of the model building process (Chatfield, 1995) and model evaluation (Mayer and Butler, 1993). By validation, we refer to the process of assessing how well a statistical procedure performs for its intended purpose, i. e. whether the method produces accurate, reliable, and meaningful results under the conditions in which it will be used (Hastie et al., 2017).

In some cases, such validation is achieved through theoretical considerations, which typically rely on assumptions and often involve asymptotic arguments. Frequently, however, validation is carried out computationally, e. g. by means of a simulation study. In the best case, such a study is based on controllable and idealized data in the following general sense: First, a ground truth exists and is known. Second, sources of noise can be influenced and, in particular, minimized. Third, the sample size can be increased arbitrarily.

Real-world data generally differs from such requirements: ground truth such as true effect sizes or dependency structures is often unknown, variability is hardly controlled, and sample sizes are limited by practical constraints. Such limitations can be overcome by using synthetic data for method validation (Morris et al., 2019): being generated *in silico* under well-defined assumptions, synthetic datasets enable the systematic evaluation of statistical methods, for example by assessing the bias of estimates, robustness to violations of assumptions, calibration of models, or sensitivity of results to different dependency structures. Consequently, synthetic data serve as a flexible and powerful tool for validating statistical methods, especially in settings where real-world datasets are unavailable, incomplete, or insufficient for rigorous performance assessment. Morris et al. (2019) present a tutorial on planning, executing and reporting simulation studies for method validation.

The use of synthetic data for statistical method validation is relevant across many applied disciplines. In medical research in particular, it has gained increasing importance, for example if datasets contain only few observations due to rare diseases or small populations, or due to measurement cost. Moreover, medical data may lack the required quality for method validation as a result of insufficient documentation, substantial amounts of missing values or measurement error, caused by time constraints in clinical routine, or inconsistent measurement intervals (Weiskopf and Weng, 2013). Although data imputation can be a viable solution against missing values, model misspecification or incompatibility can introduce bias and misestimate uncertainty (Little and Rubin, 2019, Sterne et al., 2009). In addition, privacy and data protection issues present major challenges in medical data, such that it is often publicly inaccessible or highly anonymized, consequently lacking relevant information.

Due to the substantial need for synthetic data in medical research, several approaches to generate such data have been presented. Specifically, synthetic data has been generated for specific diseases, such as diabetes (Liu et al., 2020), liver diseases (Wang et al., 2021) and depression (Du et al., 2017). An overview about synthetic data in the health care domain can be found in Murtaza et al. (2023), Giuffré and Shung (2023) or Gonzales et al. (2023).

When considering specific applications, the requirements for synthetic data become more concrete, extending beyond the general demands of knowledge of the ground truth, controllability of noise, and sample size scalability. In many contexts, it is not expected that synthetic data is medically correct in every detail in the sense that medical conclusions can be drawn from it; rather, the data should serve the purpose of the specific simulation study, namely, the validation of a statistical method for a particular research question (Morris et al., 2019). If a research question addresses the relationship between clinical patient characteristics and a diagnosis, one would choose a statistical method capable of analyzing such relationships. Accordingly, this relationship should be represented in the synthetic data and be controllable during data generation, ranging from none to moderate to strong association. In this way, the method can be validated and subsequently applied to real data with the confidence that it can be relied upon.

In this work, we consider a medical context that involves specific dependencies within the data: namely, the holistic examination of patients in whom multiple diseases may occur simultaneously and overall form a joint clinical picture. Thus, while the aforementioned studies aim to produce medical correct synthetic data for specific diseases, we seek to generate synthetic data that mirrors the interdependency between disease profiles, connected to patient information. The motivation for this research focus arises from the fact that, in reality, patients frequently suffer from several co-occurring diseases (of varying severity). These diseases present in similar, different or even antagonistic symptoms. This is particularly relevant as, in practice, establishing one diagnosis may lead to the premature exclusion of an alterative one. Such failure to recognize a co-occuring disease can have serious consequences. Examples for known co-occurring diseases with similar symptoms are diabetes mellitus and cardiovascular diseases (Siam et al., 2024), depression and anxiety disorders (Hirschfeld, 2001) or the fibromyalgia syndrome (FMS) and irritable bowel syndrome (Garofalo et al., 2023).

Our motivation for examining co-occurring diseases arose from a project in which we were concerned with the diagnosis of patients suffering from chronic pain. Such patients are often affected by multiple co-occuring diseases. Thus, we analyzed patient data with the aim to investigate whether the predictive accuracy of a disease of interest could be improved by joint consideration of comorbidities. We compared the prediction performance of two competing statistical methods: single-label classification and multi-label classification (Schmiegel et al., 2025). Our analysis provided answers to the specific research question, namely, that the inclusion of additional diagnoses did not yield substantial knowledge gain in this particular case. However, based on the real data, it was impossible to investigate under which conditions (in terms of the true underlying effect sizes, as well as data quality and sample size) one classification approach would have outperformed the other. We consider such an assessment to be highly relevant and therefore aim to conduct a simulation study using synthetic data. The present work lays the foundation for this.

Practical challenges in conducting simulation studies and, consequently, in finding or generating synthetic data for a specific research question lie in various aspects and also depend on the researcher’s level of domain-specific expertise: With regard to the medical background, relevant relationships must be appropriately identified and described. These relationships need to be statistically modeled, often by specifying a data-generating process, with particular attention typically given to conditional and unconditional as well as joint and marginal distributions. Even when such a data-generating process is known, its exact simulation often requires the use of approximate algorithms. Finally, the procedures need to be implemented using statistical software.

Ideally, suitable tools and software packages would be available for straightforward simulation of synthetic data. However, the diversity of application-specific requirements makes it challenging to develop universally applicable tools: each comes with a different focus and its own strengths. For example, the software `synthea` (Walonoski et al., 2018) and the R package `synthpop` (Nowok et al., 2016) aim to generate biologically correct data by building on real-data input. Others, such as the R packages `SimCorMultRes` (Touloumis, 2016),`bindata` (Leisch et al., 2024),`simdata` (Kammer, 2024) and`copula` (Hofert et al., 2025), operate without the need for real data. Regarding the context of our medical and methodological research question, we were unable to find a flexible framework that met our requirements for generating synthetic data for co-occurring diseases.

In our work, we develop, describe and evaluate a process for generating synthetic medical data with the aim to support statistical method validation in the context of co-occurring diseases. Specifically, we design a four-step framework, with each step building on the results from the previous one. Within each step, we propose alternative approaches that can be combined across the different steps in a modular manner.

The remainder of this paper is organized as follows: In Section 2, we formulate the statistical model of interest and guide the reader through the aforementioned four-step framework. Further, we present strategies for verifying required key properties in the synthetic data. In Section 3, we turn to the medical use case of co-occurring pain-causing diseases which provided the motivation for the methodological question. For demonstration purposes, we utilize real data as a basis for synthetic data generation, applying a selection of presented approaches. Subsequently, we examine to what extent the intended properties can be verified. In Section 4, we discuss strengths and limitations of the generation approaches and scenarios in which they are most applicable. We conclude our work in Section 5. Additional calculations and results are provided in the appendix.

## 2 Methods

We aim to provide controlled datasets that represent a medical use case and enable us, through controllable parameters, to perform method validation by means of simulation studies. The particular application of interest concerns the situation where patients suffer from multiple diseases simultaneously, and where a holistic view of patient characteristics and comorbidities can support the diagnostic process. Before addressing this specific application, we first explain the general approach to synthetic data generation.

For this purpose, one distinguishes between three types of synthetic data generation processes, each of which is suited to different research questions and the prevailing conditions. In all cases, we assume the existence of ground truth, that is, we specify characteristics of individuals and/or populations and require the synthetic data to resemble these properties. The three types are *knowledge-driven, data-driven* and *hybrid* data generation (Murtaza et al., 2023). The knowledge-driven approach uses available information from scientific literature, other publication sources and expert knowledge to describe the theoretical basis for data generation. As an example, one may sample random variables from distributions motivated by expert knowledge, or using literature-derived parameters. The data-driven approach, in contrast, is solely based on real data as input for the data generating process. Examples are parametric or nonparametric bootstrap. Eventually, the hybrid approach is a combination of defining rules, models and algorithms and incorporating real data. All three approaches come with strengths and challenges, often connected to the quality of both the domain knowledge and the input data. A detailed discussion is presented by Murtaza et al. (2023).

Apart from this categorization of the generation process, there is also a categorization of the resulting data: According to McLachlan et al. (2019), one distinguishes between *truly synthetic, fully synthetic, partially synthetic* and *anonymized-only* data. For truly synthetic data, no real data points are considered during the generation process at all. For fully synthetic data, in contrast, information from real data is used to generate new data; however, the real data points will not be part of the generated data. Partially synthetic data combines (truly or fully) synthetic data with unmodified real data. Eventually, anonymized-only data purely consists of real data, however, with sensitive information removed, replaced or encrypted. In our work, we pursue knowledge- or data-driven generation of truly or fully synthetic data.

### 2.1 Model

We are interested in the medical use case where patients suffer from several diseases and a variety of medical symptoms at the time. This may include, on the one hand, cases where different diseases exhibit similar symptoms and, as a result, a particular disease may be overlooked. On the other hand, it may also concern situations where symptoms are opposing, and the symptoms of one disease lead to the exclusion of another diagnosis.

In terms of statistical modeling, we consider a set of patients *i* = 1, …, *n*. Each patient suffers from one, several or none of *d* diseases of interest. The presence or absence of diseases is expressed by ***y***_*i*_ = (*y*_*i*1_, …, *y*_*id*_)^*′*^ ∈ {0, 1}^*d*^, where *y*_*ij*_ equals one if patient *i* suffers from disease *j* ∈ {1, …, *d*}, and zero otherwise. Additionally, there is patient information available, such as core data, symptoms or laboratory values. This is summarized in a *k*-dimensional vector ***x***_*i*_ for patient *i*, comprising values of *k* covariates.

While ***y***_*i*_ denotes the (in practice unknown) ground truth, we use ***Y*** _*i*_ = (*Y*_*i*1_, …, *Y*_*id*_)^*′*^ to denote a *d*-dimensional random variable with state space {0, 1}^*d*^, which ***y***_*i*_ is a realization of. Disease probabilities are given by

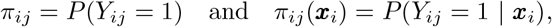

respectively. We also consider disease scores *s*_*ij*_ which have the meaning that

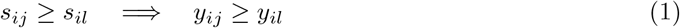

for diseases *j, l* ∈ {1, …, *d*}. Other than probabilities, they determine disease outcomes without further randomness (more details provided in Approaches IIIc, IVc and IVd below). We summarize the probabilities and scores to vectors ***π***_*i*_ = (*π*_*i*1_, …, *π*_*id*_)^*′*^ and ***s***_*i*_ = (*s*_*i*1_, …, *s*_*id*_)^*′*^, respectively.

When we speak of co-occurring diseases, particular attention is given to possible medical causes of such co-occurrences. In general, diseases may or may not have causal effect on each other; but even if they do not, their occurrence may be correlated due to covariates that are associated to both diseases. That is, the covariates may indicate the presence or absence of diseases without influencing their occurrence, or they may directly promote or prevent the occurrence of diseases, or both. In our study, we do not make any explicit assumption about the unconditional (in-)dependence of any two diseases *j* and *l*, i. e. about the joint distribution of *Y*_*ij*_ and *Y*_*il*_ for diseases *j, l* ∈ {1, …, *d*} with *j* ≠ *l*.

### 2.2 Synthetic data generation

We aim to generate synthetic data for the above described model of co-occurring diseases. We set the number of patients to *n* ≥ 1, the number of considered diseases to *d* ≥ 2 and the number of patient covariates to *k* ≥ 1. In what follows, we assume all patients (more precisely, the occurrence of their diseases and the values of their covariates) to be mutually independent and use a generic index *i* ∈ {1, …, *n*} to refer to patient *i*.

To generate medically realistic or statistically interesting data, a modeler should be able to specify certain target parameters. These include:

i. the effect of patient covariates on disease occurrence;
ii. the dependencies among the several diseases;
iii. as well as the prevalence *p*_*j*_ of individual diseases or disease combinations *j*.

It will not always be possible to satisfy every target simultaneously. For example, if two diseases are influenced by covariates in opposite ways, a positive correlation between them may be precluded. Similarly, commonly observed covariate values are less likely to be associated with rare diseases. This section describes algorithms that incorporate mathematical constraints, such as the requirement of positive definiteness for correlation matrices, and demonstrates how data can be obtained that matches the specified targets as closely as possible.

For the data generation, we start by (I) first simulating covariate data ***x***_*i*_. Then, conditional on this, we (II) construct functions which mirror the impact of covariates on disease occurrence. These are based on linear combinations of covariate values; we call them (linear) predictors and denote them by ***η***_*i*_|***x***_*i*_. Next, we (III) derive disease probabilities ***π***_*i*_|***x***_*i*_ or disease scores ***s***_*i*_|***x***_*i*_ (usually equivalent to ***π***_*i*_|***η***_*i*_ and ***s***_*i*_|***η***_*i*_). Finally, we (IV) draw realizations ***y***_*i*_|***π***_*i*_ or ***y***_*i*_|***s***_*i*_ of disease occurrences. In this last step we assume that the probabilities ***π***_*i*_ and scores ***s***_*i*_ contain sufficient information for the simulation of disease occurrences in the sense that the covariates ***x***_*i*_ do not contain any additional relevant information anymore. That is, conditional on ***π***_*i*_ or ***s***_*i*_, ***y***_*i*_ is independent of ***x***_*i*_. In the following, we describe these Steps I to IV, and within each step, a number of alternative approaches. Figure 1 provides an overview of possible combinations.

**Figure 1.**
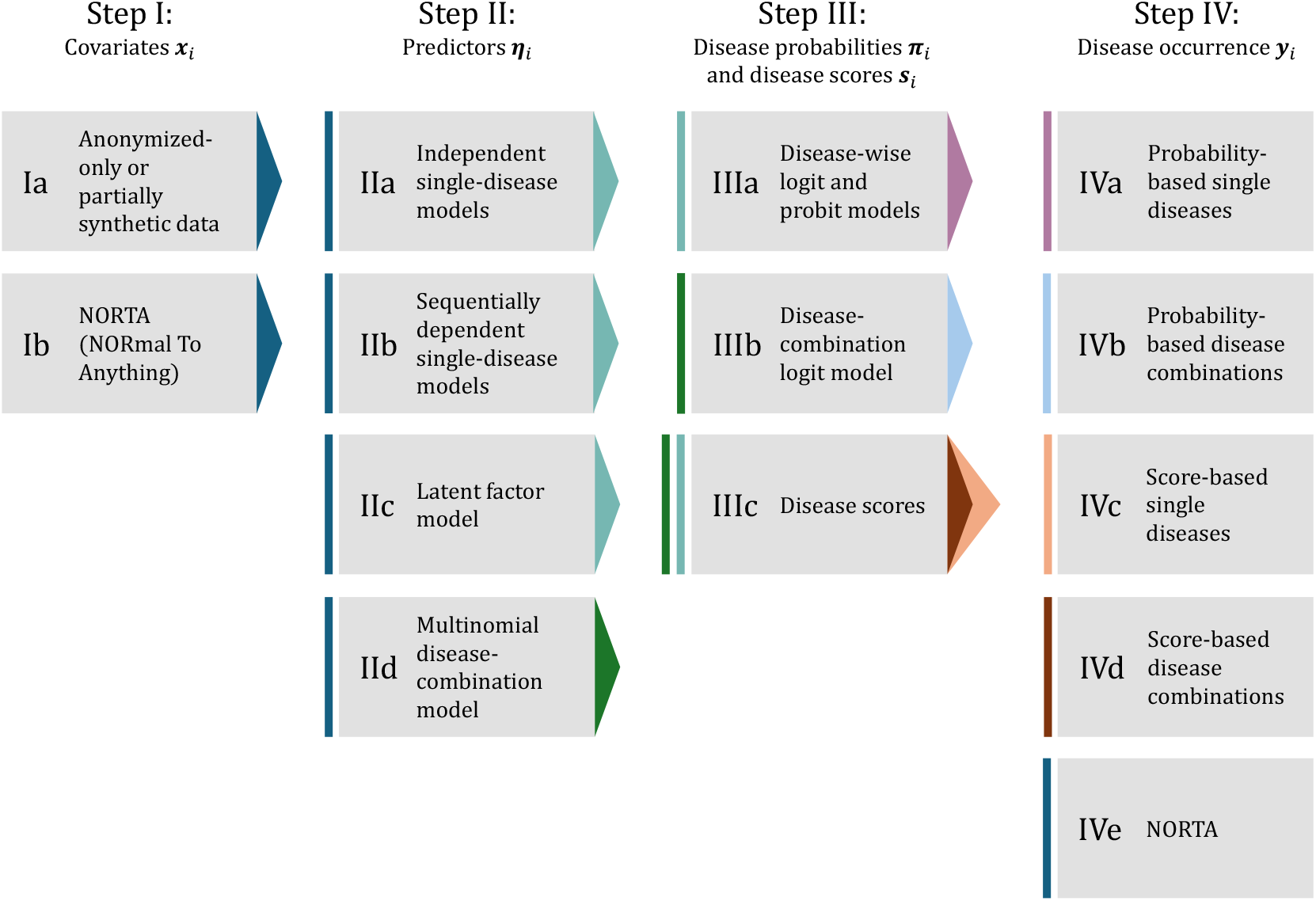
: Overview of Steps I to IV for synthetic data generation. Each column shows various approaches, representing alternative options that can be combined across steps. The colors of arrows at the right side of each box indicate which approach can follow next, namely one with a bar with matching color at the left. Note that Approach IVe can follow directly after Step I, meaning that Steps II and III are not required here.

#### 2.2.1 Covariates *x*_*i*_

Covariates capture patient details such as core data, lifestyle factors, clinical measurements, laboratory results, or genetic data. They do not yet include diagnoses but may still offer indications of potential diseases. The distribution of individual covariates, as well as the dependencies between them, are considered representative of the population of interest. Distributions should therefore be chosen accordingly, and results interpreted in this context.

##### Ia. Anonymized-only or partially synthetic data

If suitable data is available, and if data quality, data quantity, and data protection do not pose any obstacles, anonymized-only or partially synthetic data can be used as covariates, even if no corresponding diagnoses are known.

##### Ib. NORTA (NORmal To Anything)

In the following, we focus on the more complex case in which we aim for truly synthetic or fully synthetic covariates. In our approach, we create synthetic data by the NORmal To Anything (NORTA) approach (Cario and Nelson, 1997). This approach requires the modeler to specify the correlation structure and the marginal distributions of the covariates. The information basis for this specification determines whether the resulting data is regarded truly or fully synthetic. Based on Gaussian copulas, the algorithm leads to the multivariate generation of data with the desired properties, i. e. association structure and distribution. The steps are as follows:

1. Specify the correlation structure of the *k* covariates by a (*k* × *k*)-dimensional matrix ***R*** that contains all pairwise correlations. The matrix must, of course, satisfy the properties of a correlation matrix; that is, it must be symmetric, positive semi-definite, have values in the range [−1, 1] and ones on the main diagonal.
2. Describe the marginal distributions of the covariates *h* ∈ {1, …, *k*} by defining their cumulative distribution functions *F*_*h*_.
3. Incorporate the desired dependence structure between the covariates by drawing a random variable ***z***_*i*_ from a multivariate normal distribution with mean zero and covariance matrix ***R*** for each patient *i*, i. e. ***z***_*i*_ = (*z*_*i*1_, …, *z*_*ik*_)^*′*^ ∼ 𝒩 _*k*_(**0, *R***) independently for all *i* = 1, …, *n*.
4. Transform the ***z***_*i*_ into (correlated) uniformly distributed variables in the domain [0, 1]^*k*^ by component-wise application of the standard normal cumulative distribution function Φ. This way, we generate random variables ***u***_*i*_ = (*u*_*i*1_, …, *u*_*ik*_)^*′*^ with *u*_*ih*_ = Φ(*z*_*ih*_) for *i* ∈ { 1, …, *n*} and *h* ∈ { 1, …, *k*}. Each of these variables is marginally uniform, while the sought dependence structure between the variables remains.
5. Convert the uniform variables ***u***_*i*_ into the desired target distribution using the quantile functions (inverse cumulative distribution functions) 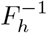 of the respective covariates by setting ***x***_*i*_ = (*x*_*i*1_, …, *x*_*ik*_)^*′*^ with 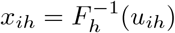 for all *i* ∈ {1, …, *n*} and *h* ∈ {1, …, *k*}.

### 2.2.2 Predictors *η*_*i*_

The predictors represent the central mechanism for establishing the association between covariates and disease occurrence, and the relationship between diseases. We present four different approaches that differ in the dependencies they generate and in their complexity, in terms of the parameters that need to be specified: independent single-disease models, sequentially dependent single-disease models, a latent factor model, and a multinomial disease-combination model. Table A1 summarizes required input specifications for these approaches.

Although mathematically not required, we recommend standardizing metric covariates *h* such that the empirical mean over *x*_1*h*_, …, *x*_*nh*_ equals zero and the empirical variance becomes one, or that they cover identical value ranges. This leads to better comparability of the effect sizes to be specified. In the following, all computations of predictors ***η***_*i*_ are conditional on ***x***_*i*_, even if not flagged explicitly in equations. We employ vectors 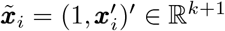 for the purpose of vector product notation.

#### IIa. Independent single-disease models

We use regression models (Fahrmeir et al., 2013) to describe the probabilities of patients suffering from the considered diseases, conditional on the covariate information. To that end, we start by combining the covariates to linear predictors. In our first approach, we model the diseases to occur independently of each other. In particular, we do not assume that diseases are mutually exclusive; instead, a patient can suffer from any combination of them. In detail, we follow these steps for patients *i* ∈ {1, …, *n*} and diseases *j* ∈ {1, …, *d*}:

- Define coefficient vectors ***β***_*j*_ = (*β*_*j*0_, …, *β*_*jk*_)^*′*^, where *β*_*jh*_ ∈ ℝ describes the association between the *j*th disease and *h*th covariate, conditional on all others. The desired proportion *p*_*j*_ of patients affected by disease *j* can be regulated by the intercept *β*_*j*0_. In case of normally distributed covariates with mean zero, it should be chosen as

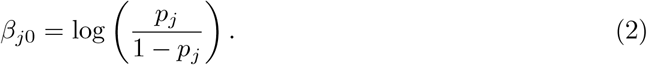
- Calculate a linear predictor *η*_*ij*_ | ***x***_*i*_ as

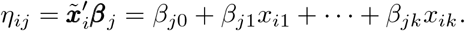

#### IIb. Sequentially dependent single-disease models

Next, we extend Approach IIa by using sequentially dependent rather than independent regression models to describe disease probabilities. This modeling approach is based on the assumption that information about the presence of one disease alters the probability of another disease being present, beyond what is already explained by the information contained in the covariates. To include disease dependencies in the simulation process, we model the disease occurrences sequentially and condition on the already generated disease variables when calculating the linear predictor for subsequent diseases. To this end, we follow these steps for every patient *i* ∈ {1, …, *n*}:

1. Define coefficient vectors ***β***_*j*_ = (*β*_*j*0_, …, *β*_*jk*_)^*′*^ as in Step 1 of Approach IIa. Note, however, that the coefficients will have a different impact here due to the conditioning on other diseases (inclusion of coefficients *γ*_*jl*_) in the following.
2. Sort the diseases to obtain an order such that, for *j* < *l*, we can model the probability of occurrence of disease *j* independently of disease *l*. In particular, the occurrence probability of disease 1 will be modeled independently from all other diseases.
3. Determine coefficients *γ*_*jl*_ which describe how the occurrence probability of disease *l* is influenced by the occurrence of disease *j* ∈ {1, …, *l* − 1}, conditional on ***x***_*i*_ and the occurrence information of the diseases 1, …, *j* − 1, *j* + 1, …, *l* − 1.
4. For *l* = 1, do the following:
  - Calculate

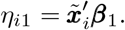
  - Proceed with Steps III and IV to simulate *y*_*i*1_.
5. For *l* = 2, …, *d*, do the following:
  - Calculate

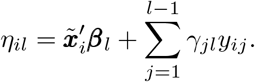
  - Proceed with Steps III and IV to simulate *y*_*il*_.

#### IIc. Latent factor model

So far, we have computed the predictors conditional on observed covariates and (in Approach IIb) information about the presence of other diseases. In a further approach, we now include additional factors that are neither observed nor assigned a substantive interpretation. These are latent factors that influence the joint occurrence of each pair of diseases. Using shared latent factors makes it possible to simulate disease occurrences which depend on both known patient covariates and unknown other influences (Greene, 2019). We proceed as follows:

1. Define coefficient vectors ***β***_*j*_ = (*β*_*j*0_, …, *β*_*jk*_)^*′*^ as in Step 1 of Approach IIa. Note, however, that the coefficients will have a different impact here due to the conditioning on latent factors (inclusion of coefficients *λ*_*jl*_) in the following.
2. For every disease *j*, determine a set *B*_*j*_ of diseases which share a latent factor with *j*, i. e. *B*_*j*_ ∈ {1, …, *d*} *\* { *j*}. By definition, shared latent factors influence two diseases; thus, the sets must fulfill

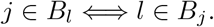

It is allowed that *B*_*j*_ is empty. To avoid using vectors of varying dimensions in the following, we introduce the (symmetric and potentially sparse) matrix ***B***^∗^ with entries *b*_*jl*_ = 1 if *j* ∈*B*_*l*_, i.e. if diseases *j* and *l* share a latent factor, and zero otherwise.

- Define coefficients *λ*_*jl*_ ∈ ℝ which indicate the influence of the latent factor shared by diseases *j* and *l* on disease *j*. Although the latent factor is shared, the influence on diseases *j* and *l* may differ, i. e. we do not require *λ*_*jl*_ = *λ*_*lj*_. For *b*_*jl*_ = 0, we set *λ*_*jl*_ = 0.
- For each patient *i*, simulate independent standard normally distributed latent factors *w*_*ijl*_ ∼ *N* (0, 1) if *b*_*jl*_ = 1 and zero otherwise. Since latent factors are shared, we require *w*_*ijl*_ = *w*_*ilj*_ for all *j, l* ∈ {1, …, *d*}.
- For each patient *i* and disease *j*, calculate a linear predictor which takes both the influence of covariates ***x***_*i*_ as well as latent factors *w*_*ij*1_, …, *w*_*ijd*_ into account:

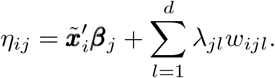

#### IId. Multinomial disease-combination model

So far, we have considered the *d* diseases side by side and, for each patient, examined a *d*-dimensional combination (*y*_*i*1_, …, *y*_*id*_)^*′*^ ∈ {0, 1}^*d*^ of occurrences and non-occurrences. An alternative approach is to describe each combination as one of 2^*d*^ possible (unordered) classes of aggregated disease profiles. This is what we do in the following. To that end, we introduce variables *c*_*i*_ with *c*_*i*_ = *r* indicating that patient *i* suffers from disease combination *r*, and *r* ∈ {1, …, 2^*d*^}. In particular, we number the classes such that combination (*j*_1_, …, *j*_*d*_)^*′*^ ∈ {0, 1}^*d*^ corresponds to class 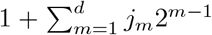. For example, in the case of three possible diseases, the combination (0, 0, 0)^*′*^ corresponds to class 1, (0, 1, 0)^*′*^ to class 3, and (1, 1, 1)^*′*^ to class 8. For any *d*, the first class refers to the combination where none of the single diseases occurs, and *r* = 2^*d*^ to the case where all of them do. We undertake the following steps:

1. For every disease combination *r*, we calculate linear predictors after defining regression coefficients as intercepts and the effects of the *k* covariates on the disease outcome. For patient *i*, these linear predictors *η*_*ir*_ represent the log-odds of class *r* relative to the reference category. Since *r* = 1 serves as reference category and its occurrence probability will later (in Step III) be computed as the complement to all other class probabilities, we do not specify coefficients for this class.

We propose three ways to define the regression coefficients and linear predictors:

i. If knowledge is available about occurrences of disease combinations, and how they are influenced by the covariates, the regression coefficients ***β***_*r*_ = (*β*_*r*0_, *β*_*r*1_, …, *β*_*rk*_)^*′*^ can be chosen individually for every disease combination *r*. This procedure requires the user to define (2^*d*^ − 1)(*k* + 1) coefficients *β*_*rh*_ (*r* ∈ {2, …, 2^*d*^}, *h* ∈ {0, …, *k*}).
In case of normally distributed covariates with mean zero, the intercept *β*_*r*0_ should be chosen as

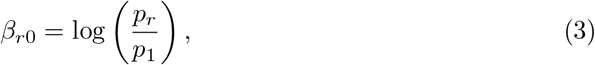

where *p*_*r*_ is the desired proportion of patients affected by disease combination *r*. This regulates the proportion of patients suffering from *r* ≥ 2 compared to the proportion of healthy patients (*r* = 1).
For every patient *i*, calculate the linear predictor for disease combination *r* ≥ 2 as

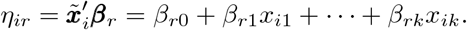
ii. Alternatively, we can model the impact of covariates on disease occurrence for every disease individually. That means, the regression coefficients are independently defined as ***β***_*j*_ = (*β*_*j*0_, *β*_*j*1_, …, *β*_*jk*_)^*′*^ for every disease *j* ∈ {1, …, *d*}. This requires the user to specify only *d*(*k* + 1) coefficients *β*_*jh*_ (*j* ∈ {1, …, *d*}, *h* ∈ {0, …, *k*}).
For every patient *i*, one calculates a linear predictor for disease *j* as

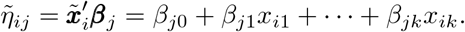

The linear predictor for disease combination *r*, corresponding to (*j*_1_, …, *j*_*d*_)^*′*^, then results as

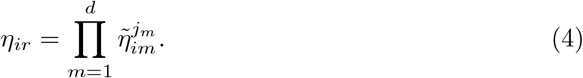

Since all exponents equal either zero or one, that is the product of all linear predictors corresponding to the single diseases in combination *r*.
iii. Via the product (4), the previous approach mingles individual linear predictors and thus eventually leads to synthetic disease occurrences which are dependent both among each other and on the covariates. However, it does not retain the individual relationship between single diseases and covariates. This can be achieved if the linear predictor for a disease combination is calculated dependent on the linear predictors of a subset of the disease combinations which reflect mutually exclusive single diseases (e. g. (1, 0)^*′*^ and (0, 1)^*′*^ for *d* = 2). To this end, we make use of the odds ratio OR_*ir*_, which quantifies for patient *i* the association between the single diseases contained in combination *r*, and therefore how the presence or absence of one single disease changes the odds of observing a particular disease category. With the detailed calculations in Appendix A.1, we show the following: Let *r* = (*j*_1_, …, *j*_*d*_)^*′*^ be some disease combination with *j*_1_ = 1 and *j*_2_ = 1 (and arbitrary values for *j*_3_, …, *j*_*d*_. Let *r*^*′*^ be identical to *r* with the difference that *j*_1_ = 0, and *r*^*′′*^ like *r* with *j*_2_ = 0. Then, the linear predictor for patient *i* and disease combination *r* can be calculated as:

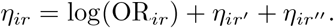

We can set OR_*ir*_ according to the desired association between the diseases 1 and 2. It then holds that

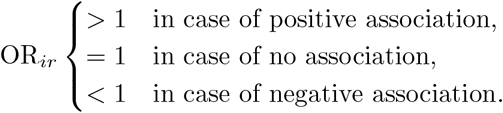

With this procedure, we constrain the linear predictor for the joint disease outcome instead of building independent linear predictors. This allows us to explicitly model an association between the single diseases. In Section A.1, we also present the calculations for a combination of three diseases. The derivation for more than three diseases is analogous; however, combining more diseases decreases their practical relevance.

### 2.2.3 Disease probabilities *π*_*i*_ and disease scores *s*_*i*_

After having constructed predictors that link the values of covariates with the occurrence of single or combined diseases, we now aim to derive probabilities ***π***_*i*_ or scores ***s***_*i*_ for disease occurrences based on them. In the following, we use the common notation to condition on the covariates ***x***_*i*_ rather than on the predictors ***η***_*i*_, because the relation between ***x***_*i*_ and ***η***_*i*_ is often deterministic. Probabilities have the advantage of being clearly interpretable, with a known range and an intuitive sense of how common or rare events are, and they directly represent the uncertainty associated with random events. Depending on the context, however, one may instead be interested in scores, which preserve the structure of the (linear) predictors and allow for easier control of the simulated disease occurrences. In the following, we present methods for deriving both: Approaches IIIa and IIIb for probabilities, and IIIc for scores.

#### IIIa. Disease-wise logit and probit models

Approaches IIa, IIb and IIc delivered predictors *η*_*ij*_ for patients *i* ∈ {1, …, *n*} for every single disease *j* ∈ {1, …, *d*}. Dependencies between diseases are integrated in the predictors already. A straightforward procedure from regression modeling is to use the logistic function to translate the predictors into occurrence probabilities

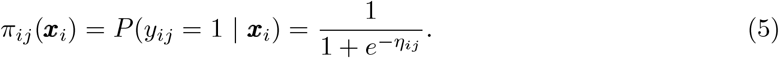

Alternatively, corresponding to the well-known probit model, one may calculate

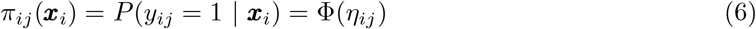

with the cumulative distribution function Φ of the standard normal distribution. While the logit model (5) is generally more straightforward to interpret, the probit model (6) comes with properties such as thinner tails.

#### IIIb. Disease-combination logit model

Approach IId considered disease combinations (*y*_*i*1_, …, *y*_*id*_)^*′*^ ∈ {0, 1}^*d*^, numbered by *r* ∈ {1, …, 2^*d*^}. Accordingly, we derive multinomial probability vectors 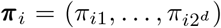. These differ from the probabilities in Approach IIIa in that the components are required to sum up to one. Based on *η*_*ir*_, we calculate the occurrence probability for disease combination *r* ≥ 2 as

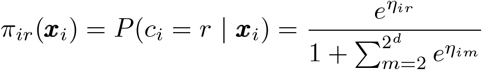

where *c*_*i*_ stands for the disease combination class of patient *i* as introduced in Approach IId. The occurrence probability for the reference category *r* = 1 can be calculated as 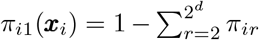 (Fahrmeir et al., 2013).

#### IIIc. Disease scores

Unlike probabilities, disease scores determine disease outcomes directly, without introducing further uncertainty, through Formula (1) for diseases or disease combinations. In the simplest case, one may leave the predictors from Step II unaltered and set *s*_*ij*_ = *η*_*ij*_ for all *i* and *j*. Alternatively, one may add independent and identically distributed noise *ε*_*ij*_ ∼ *N* (0, *σ*^2^) for some *σ* > 0, such that *s*_*ij*_ = *η*_*ij*_ + *ε*_*ij*_. The individual- and disease-specific noise term represents the fact that, even if two individuals agree in their covariates and potentially latent factors, the disease outcome may differ.

### 2.2.4 Disease occurrence *y*_*i*_

Finally, we proceed from the continuous-valued probabilities and scores to discrete-valued realizations of whether a disease or disease combination does or does not occur. While the probability-based Approaches IVa and IVb represent the uncertainty associated with such transitions more realistically, the score-based Approaches IVc and IVd offer greater controllability, particularly with respect to desired disease prevalences. Approach IVe revisits the NORTA algorithm and generates disease occurrences based on correlations.

#### IVa. Probability-based single diseases

Given we have calculated occurrence probabilities *π*_*ij*_ for individual diseases *j* (via Approaches IIIa or IIId), we now draw independent Bernoulli distributed random variables:

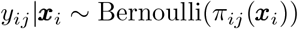

for all patients *i* ∈ {1, …, *n*} and diseases *j* ∈ {1, …, *d*}.

In Approach IIa, we indicated a choice of intercept in Equation (2) which (in theory) regulates the proportion of patients suffering from disease *j* such that it matches the desired fraction *p*_*j*_. In case of non-normally distributed or non-centered covariates, however, the target proportion may be missed. In this case, an iterative correction can be used where one adjusts the intercept *β*_*j*0_ iteratively to

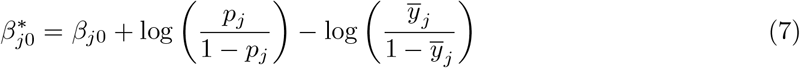

until the difference between the target prevalence *p*_*j*_ and sample proportion 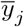 falls below a chosen tolerance value (e. g. 0.001) (King and Zeng, 2001).

#### IVb. Probability-based disease combinations

If we obtained occurrence probabilities *π*_*ir*_ for disease combinations *r* ∈ { 1, …, 2^*d*^} rather than individual diseases (via Approach IIIb), we proceed by drawing independent multinomial distributed random variables:

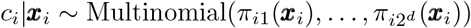

for all patients *i* ∈ {1, …, *n*}, where *c*_*i*_ denotes the disease combination for patient *i*. Before, we numbered the classes such that combination (*j*_1_, …, *j*_*d*_)^*′*^ corresponded to class 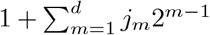. We can thus determine the individual disease occurrences for patient *i* via

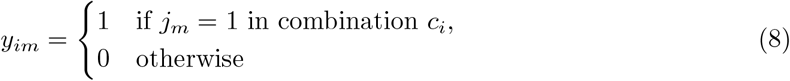

for *m* ∈ {1, …, *d*}.

In analogy to the intercept correction in Equation (7) from Approach IVa, the intercept *β*_*r*0_ for disease combination *r* ≥ 2 can be iteratively adjusted by

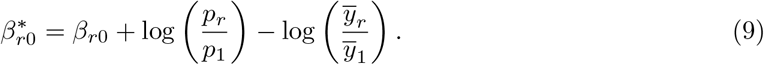

#### IVc. Score-based single diseases

Based on disease scores *s*_*ij*_ for single diseases *j* ∈ {1, …, *d*}, one can determine binary disease outcomes by comparing the scores with thresholds *τ*_*j*_ which are chosen by the modeler for every disease *j*:

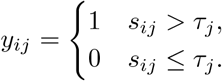

Adjusting the threshold *τ*_*j*_ enables one to reach the desired prevalence *p*_*j*_ of disease *j*. In particular, one can chose *τ*_*j*_ to be the empirical *p*_*j*_-quantile across *s*_1*j*_, …, *s*_*nj*_.

#### IVd. Score-based disease combinations

If scores *s*_*ir*_ for disease combinations *r* ∈ {1, …, 2^*d*^} are provided, a combination can be selected as

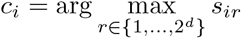

for every patient *i* ∈ {1, …, *n*}. If the score was obtained by adding normally distributed noise in Approach IIIc, this prodecure corresponds to a multivariate probit model. Via Equation (8), one can obtain single-disease realizations from the combinations.

#### IVe. NORTA

In this approach, we utilize the NORTA procedure which has already been described in Approach Ib in the context of generating synthetic covariate data. It directly builds on the covariate generation (Step I) without taking the model-based path through Steps II and III: It simulates disease occurrence solely based on specified marginal distributions and correlations. With the following algorithm, we extend the framework of Cario and Nelson (1997). In particular, we describe how occurrences *y*_*ij*_ are generated conditional on covariates ***x***_*i*_. We provide this description according to the chosen structuring into Steps I through IV. However, if NORTA was already applied in Step I (i. e. Approach Ib), these steps could be merged for easier implementation, without the need to consider the subsequent formulas for conditional probabilities. If covariate data was obtained via Approach Ia, or if one seeks to separate the levels, one should follow these steps:

1. Determine the correlation structure of the *k* covariates. This is either known, if the covariates were synthetically generated by Approach Ib, or it can be estimated via the empirical correlation matrix of the ***x***_*i*_. In any case, denote the resulting (*k × k*)-dimensional matrix by ***R***_*xx*_.
2. Specify the correlation structure of the *d* diseases (i. e. (*y*_*i*1_, …, *y*_*id*_)^*′*^ across all patients *i*) by a (*d × d*)-dimensional matrix ***R***_*yy*_ that contains all pairwise correlations. Further, specify the pairwise correlations between (*y*_*i*1_, …, *y*_*id*_)^*′*^ and (*x*_*i*1_, …, *x*_*ik*_)^*′*^ across all *i* in a (*d × k*)-dimensional matrix ***R***_*yx*_.
3. Describe the marginal distributions of the occurrences *y*_*ij*_ by defining their cumulative distribution functions *F*_*y,j*_ for *j* ∈ {1, …, *d*}.
4. Generate auxiliary random vectors ***z***_*x,i*_ which correspond to the vectors ***z***_*i*_ in Step 3 of Approach Ib. If these vectors are still known from a previous NORTA application, they can be used. Otherwise, conduct the following calculations (reversal of Steps 4 and 5 from Approach Ib) based on the available covariate data:
  - Compute the empirical cumulative distribution functions *F*_*x,h*_ for all covariates *h*.
  - Calculate *u*_*x,ih*_ = *F*_*x,h*_(*x*_*ih*_) and *z*_*x,ih*_ = Φ^−1^(*u*_*x,ih*_) for all *i* and *h*, where Φ denotes the standard normal cumulative distribution function. Let ***z***_*x,i*_ = (*z*_*x,i*1_, …, *z*_*x,ik*_)^*′*^.
5. Incorporate the desired dependence structure among the diseases and between the diseases and covariates by drawing random variables ***z***_*y,i*_ = (*z*_*y,i*1_, …, *z*_*y,id*_)^*′*^ from a multivariate normal distribution with mean

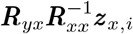

and covariance matrix

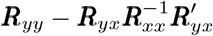

independently for all patients *i* ∈ {1, …, *n*}.
6. Transform the ***z***_*y,i*_ into (correlated) uniformly distributed variables in the domain [0, 1]^*d*^ by component-wise application of Φ. This way, we generate variables ***u***_*y,i*_ = (*u*_*y,i*1_, …, *u*_*y,id*_)^*′*^ with *u*_*y,ij*_ = Φ(*z*_*y,ij*_) for *i* ∈ {1, …, *n*} and *j* ∈ {1, …, *d*}. Each of these variables is marginally uniform, while the sought dependence structure between the variables remains.
7. Convert the uniform variables ***u***_*y,i*_ into the desired target distribution using the quantile functions (inverse cumulative distribution functions) 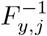 of the respective occurrences by setting *y*_*ij*_ = *F* ^−1^(*u*_*y,ij*_) for all 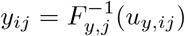 and *j* ∈ {1, …, *d*}.

The NORTA approach can also be used to generate occurrences of disease combinations rather than single diseases.

### 2.3 Verification of desired properties

In the beginning of Section 2.2, we outlined three target properties which we aim to fulfill with the generated data: (i) covariate effects on disease occurrence, (ii) associations between diseases, and (iii) disease prevalences. Once the synthetic data has been generated using the above described approaches, we seek to assess to what extent it reflects the desired ground truth. We proceed as follows:

i. **Covariate effects:** The connection between patient covariates and disease occurrence was established in Step II through the construction of predictors. In the approaches presented, regression coefficients were defined to combine the covariates into linear combinations. Due to the superposition of covariate effects with inter-disease dependencies, it can, however, be assumed that the original relationship has become biased. We estimate the coefficients of regression models to verify the consistency between the input parameters and the reverse-engineered estimates. To that end, we look at different paths to generate data, and apply five different ways to re-estimate the input parameters: independent binomial logit models, dependent binomial logit models, a probit model, a multinomial logit model, and the computation of empirical correlations.
ii. **Association between diseases:** To examine the relationship between the generated disease occurrences, we use Cramer’s V (Cramér, 1946), which is a measure of the association strength between two nominal variables. The measure ranges between zero and one, where *V* = 0 means no association and *V* = 1 means perfect association between the according variables. Another meaningful measure, though not applied here for verification purposes, is the tetrachoric correlation, which measures the correlation between two binary variables for which one assumes that they arose from an underlying normally distributed latent variable (Greene, 2019).
iii. **Target prevalence:** To assess if the desired disease prevalences are reached, we compare for each disease the target proportion with the arithmetic mean of the according generated disease occurrence.

## 3 Use case: Co-occurring diseases in chronic pain patients

To test and demonstrate the practical application of the four-step framework in a real-world context, we create and evaluate synthetic datasets for the medical use case which motivated this study.

### 3.1 Medical context and data

We consider the situation of chronic pain patients who often suffer from multiple co-occurring diseases. In Schmiegel et al. (2025), we analyze data from a German rheumatism outpatient clinic, the Department of Internal Medicine and Rheumatology at Hospital Bielefeld Rosenhöhe. All patients in this dataset are suspected to suffer from none, one or several of the following diseases: immune-mediated rheumatic diseases (IMID), fibromyalgia syndrome (FMS), osteoarthritis (OA) and other pain-causing diseases (OPCD). The aim of Schmiegel et al. (2025) was to investigate whether the data-driven diagnosis of IMID can be improved by the simultaneous examination of FMS, OA and OPCD.

As a result of this project, methodological questions emerged that could be appropriately investigated on well-suited data. However, although a basis of real data is available in this use case, it cannot be employed for rigorous method validation due to the lack of controllability of ground truth, apart from limitations such as small sample size, measurement uncertainty and missing values. For that reason, we use the real dataset to extract a knowledge base which serves to specify distributions, ranges and interdependencies. To revisit the categorization introduced at the beginning of Section 2, we focus on data-driven, fully synthetic data generation.

#### 3.2 Procedure

For a focused demonstration of the data generation, we will subsequently concentrate on two of the aforementioned diseases, namely IMID and FMS. Even if we do not aim for medical correctness in this demonstration example, we seek to define realistic value ranges for the variables. Therefore, we use the real patient data for knowledge discovery (data-driven), while no real data is included in the final generated dataset (fully synthetic).

In the following, we describe the relevant design choices in applying the algorithms from Section 2. All details, including input and target values, can be found in the corresponding `R` scripts at https://github.com/fuchslab/synthData_diseases.

##### General settings and target properties

We take five methodological paths to generate synthetic data, illustrated in Table 1. With each path, we generate 1,000 independent and identically distributed synthetic datasets. In each of these datasets, we simulate data for *n* = 10, 000 patients. For each of the patients, in turn, we simulate the occurrence of *d* = 2 diseases as binary outcomes, namely of IMID and FMS. For each patient, we assume information from *k* = 6 covariates to be available.

**Table 1.** Overview of the five methodological paths that are chosen to generate synthetic data in the presented use case. For each Path A to E, the table lists the sequence of chosen approaches in Steps I to IV.

| Path | Step I | Step II | Step III | Step IV |
| --- | --- | --- | --- | --- |
| A | Ib | IIa | IIIa | IVa |
| B | Ib | IIb | IIIa | IVa |
| C | Ib | IIc | IIIa | IVa |
| D | Ib | IId | IIIb | IVb |
| E | Ib | – | – | IVe |

Regarding the target properties (covariate effects, disease dependencies, disease prevalences), we base the following values on characteristics from the real dataset:

i. Desired covariate effects are described further below.
ii. As an association between the two diseases IMID and FMS, we target at *V*_IMID, FMS_ = 0.42.
iii. Regarding marginal prevalences for disease occurrences, we aim to achieve a proportion of *p*_IMID_ = 0.52 for IMID and *p*_FMS_ = 0.21 for FMS. Additionally, we define the following target prevalences for the four possible disease combinations: *p*_onlyIMID_ = 0.49, *p*_onlyFMS_ = 0.19, *p*_IMID&FMS_ = 0.03 and *p*_healthy_ = 0.29.

We deliberately choose *p*_IMID&FMS_ ≠ *p*_onlyIMID_ *·3 p*_onlyFMS_, where equality would indicate marginal independence of the occurrence of the two diseases.

##### Step I: Covariates

We generate data for six patient-specific covariates: age, sex, C-reactive protein (CRP), sum of positively answered questions of the Fibromyalgia Rapid Screening Tool (FiRST), numeric rating scale (NRS) to quantify the pain level, and the number of pain-sensitive pressure points on the body (tender points). In the following, we refer to these variables as `Age`, `Sex`, `CRP`, `FiRST`, `NRS` and `Tender`. We empirically derive the distributions and their corresponding parameters from the real hospital data as listed in Table 2.

**Table 2.** Description and specification of patient variables that are generated in Step I according to distributions and parameters from real data. The simulated variables serve as covariates in Paths A to E. (*µ, σ, a, b*) 𝒯 𝒩 denotes a truncated normal distribution with mean *µ* ∈ [*a, b*], standard deviation *σ >* 0 and values being constrained to the range [*a, b*]. Γ(shape, rate) denotes the gamma distribution with positive shape and rate parameters.

| Variable | Description | Type | Distribution/Parameters |
| --- | --- | --- | --- |
| Age | Age of patient at admission (in years) | Metric (continuous) | $\mathcal{TN}(\mu = 55, \sigma = 15.5, a = 18, b = 89)$ |
| Sex | Sex at birth | Binary | Bernoulli($p = 0.31$); 1 = male, 0 = female |
| CRP | C-reactive protein (in mg/l) | Metric (continuous) | $\Gamma(\text{shape} = 0.5, \text{rate} = 0.05)$ |
| FiRST | Sum of positively answered questions of the Fibromyalgia Rapid Screening Tool | Metric (discrete) | Sample from $\{0, 1, \dots, 6\}$ with probabilities 0.03, 0.10, 0.13, 0.17, 0.19, 0.18, 0.20 |
| NRS | Numeric rating scale to quantify the pain level | Metric (discrete) | Sample from $\{0, 0.5, 1, \dots, 9.5, 10\}$ with probabilities 0.031, 0.002, 0.005, 0.002, 0.020, 0.002, 0.046, 0.009, 0.046, 0.005, 0.147, 0.016, 0.153, 0.024, 0.121, 0.021, 0.178, 0.019, 0.093, 0.004, 0.056 |
| Tender | Number of pain-sensitive pressure points on body | Metric (discrete) | Sample from $\{0, 1, \dots, 18\}$ with probabilities 0.268, 0.016, 0.156, 0.021, 0.093, 0.015, 0.077, 0.005, 0.051, 0.006, 0.053, 0.011, 0.036, 0.010, 0.029, 0.015, 0.058, 0.006, 0.074 |

We generate the covariates using Approach Ib, that is the NORTA procedure. To that end, we calculate the empirical (6 × 6)-dimensional correlation matrix of the covariates (results shown in Table A2) and pass it to the algorithm as input matrix ***R***. In particular, we proceed as follows: For pairs of variables that are both continuous in the real data, we use Pearson’s correlation coefficient (Pearson, Karl, 1896); for pairs containing one continuous and one binary variable, we calculate the polyserial correlation (Drasgow, 2004, Fox, 2022). As marginal distributions, we use the ones listed in Table 2, which are motivated through empirical analysis of the real data. For all five paths listed in Table 1, we use the exact same covariate data. We standardize the metric covariates before proceeding with Step II.

##### Step II: Predictors

We apply the four model-based Approaches IIa to IId to generate predictors, entering Paths A to D. Path E does not require the generation of a predictor since the NORTA technique in Approach IVe directly builds on Step I without visiting Steps II and III. More precisely, the covariates and occurrences are generated in a joint NORTA application, and then separated for use in Paths A to D. Approaches IIa to IId require specifying coefficients that describe the effect of covariates on disease occurrences. To obtain realistic values for these coefficients, we apply regression models to the real data and use the estimated coefficients as input values for data generation. Specific choices are as follows:

- As a basis for Approaches IIa to IIc, we estimate two logistic regression models on the real data after standardizing metric covariates — one with IMID as target variable, one with FMS. This results in coefficients ***β***_IMID_ and ***β***_FMS_ that are shown in Table A3.
- In Approach IIb, we choose the order of diseases such that FMS occurrence is simulated first, and IMID occurrence is generated conditional on the occurrence of FMS. For the latter, we employ the coefficient value *γ*_FMS, IMID_ = 4.6 to model the impact of the occurrence of FMS on IMID.
- For Approach IIc, we create latent factors *w*_*i*,IMID, FMS_ for every patient *i*, which represent a shared influence on the occurrence of both IMID and FMS. This latent factor enters the linear predictor with weights *λ*_IMID, FMS_ = *λ*_FMS, IMID_ = 5.7.
- As a basis for Approach IId, we estimate a multinomial regression model on the real data after standardizing the metric covariates, where the target variable can fall into one of the four categories ‘onlyIMID’ (patients who are exclusively diagnosed with IMID), ‘onlyFMS’ (patients who are exclusively diagnosed with FMS), ‘IMID&FMS’ (patients who are diagnosed with both IMID and FMS) and ‘healthy’ (the reference category). This yields regression coefficients ***β***_onlyIMID_, ***β***_onlyFMS_ and ***β***_IMID&FMS_ that are shown in Table A3. Within the first step of Approach IId, we choose Option (i) to build linear predictors; that is, we define regression coefficients for each of the disease combination classes (except for the reference category).

##### Step III: Probabilities and scores

To reduce the variability between datasets across paths to a few sources, we generate only probabilities in Step III and refrain from producing scores. That is, we apply Approach IIIa for the single-disease Paths A to C and Approach IIIb for the disease-combination Path D.

##### Step IV: Disease occurrence

As a logical consequence of generating probabilities in Step III along Paths A through D, these paths now lead to probability-based realizations of disease occurrences: the application of Approach IVa in the single-disease cases (Paths A to C) and of Approach IVb in the disease-combination case (Path D).

In Approaches IIa to IId, we had chosen the intercept as estimated from the real data. Here, in Approaches IVa and IVb, we apply the iterative intercept optimization, described through Equations (7) and (9), respectively, in order to achieve the target proportions of diseased patients. On Path E, we move directly from the covariate data generation in Step I to the simulation of disease occurrences via NORTA in Approach IVe. To that end, we specify all pairwise correlations between the diseases IMID and FMS, and between the two diseases and the covariates. These are empirically derived from the real data and listed in Table A2. As before, for pairs containing one continuous and one binary variable, we calculate the polyserial correlation (Drasgow, 2004, Fox, 2022). For pairs of binary variables, we calculate the tetrachoric correlation (see Section 2.3). In terms of the description of Approach IVe in Section 2.2.4, they form the matrices ***R***_*yy*_ and ***R***_*yx*_. In practice, however, we generate the covariates and occurrences in one joint NORTA application and separate them afterwards. The marginal distributions of disease occurrence are binomial with success probabilities *p*_IMID_ and *p*_FMS_ as specified before.

### 3.3 Results

The data generation process involved several steps in which different model layers overlapped and interacted. We now examine to what extent the input parameters can be reproduced from the obtained datasets, and, more importantly, to what extent the synthetic data matches the main target properties (covariate effects, associations between diseases, target prevalences). To that end, we employ the measures described in Section 2.3 (estimated regression coefficients, associations in terms of correlation coefficients and Cramer’s V, and empirical disease prevalence). By comparing the results from Paths A to E, we can capture the influence of, in particular, Approaches IIa through IId, as well as the difference between the model-based Paths A to D and the correlation-based Path E.

For all data generation paths and each characteristic of interest (regression coefficients, correlations), we report the deviation of the estimated value (which we compute from the synthetic data) from the input value (that was provided as input to the data generation process and is known to the modeler). In particular, we look at the average of relative differences across all 1,000 generated datasets per path; the relative difference is the difference between the estimated value and the input value, divided by the input value. Further, we display 95 % empirical ranges of the relative differences.

Depending on the context, however, the value of the regression coefficient or the correlation resulting from the synthetic data may not be particularly important. More crucial might be that the direction of the effect is preserved, that is, a positive association should remain positive. Therefore, we also examine whether the sign between the input and estimate is retained. Overall, the metrics allow us to assess central tendencies while accounting for variation across all datasets.

Paths A to E differ in how they establish the relationship between covariates and diseases, as well as in how co-occurrences of diseases are represented. Accordingly, the calculated difference between estimate and input will depend on which model is used for reverse-engineering. To ensure that each data generation path is treated equally, we perform the parameter estimation in five different ways: independent binomial logit models, dependent binomial logit models, a probit model, a multinomial logit model, and the computation of empirical correlations (where the type of correlation depends on the data type of each pair of variables). Figure 2 displays relative differences for the regression coefficient *β*_*j*,Age_ for all combinations of data generation paths and reverse-engineering methods. It also indicates how often the signs of the estimate and the input, i. e. the direction of the effect coincide. Regarding the employed methods, the combinations on the main diagonal match best, and thus are expected to deliver the smallest differences. These method-wise optimal combinations are thus compiled for all six covariates in Figure 3. A more detailed representation, analogous to Figure 2, for the covariates beyond `Age` are provided in Figures A1 to A5.

**Figure 2.**
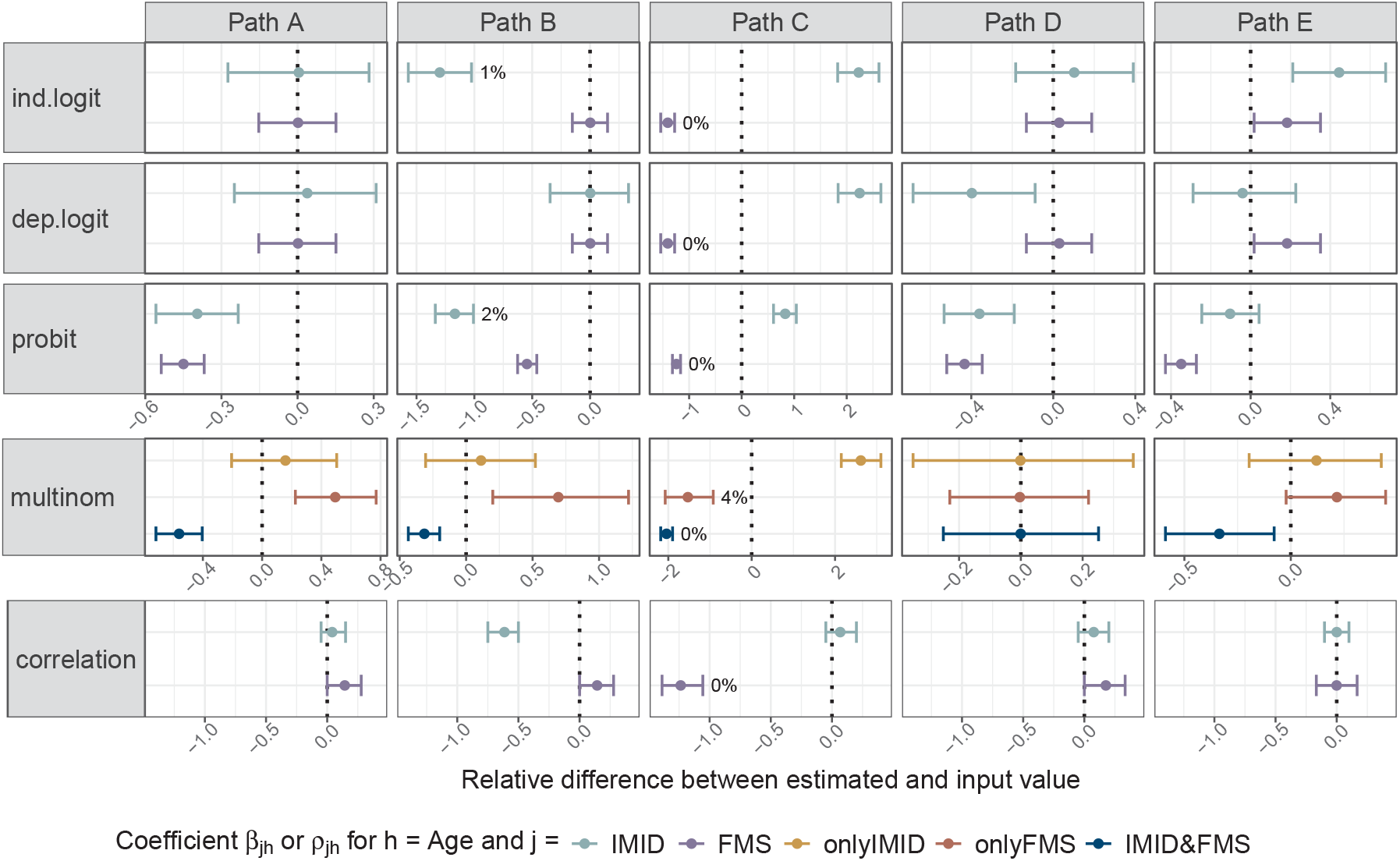
Relative difference between the estimated value of the regression coefficient *β*_*jh*_ or correlation coefficient *ρ*_*jh*_ and the input value. The plots display averages (dots) and 95 % empirical ranges (bars) across 1,000 generated datasets. Percentages indicate how often the signs of estimate and input coincide. Where no percentage is given, it is 100 %. Columns represent the data generating approach (Paths A to E), while rows represent ways to estimate *β*_*jh*_ or *ρ*_*jh*_, respectively. These are independent binomial logit models (‘ind.logit’), dependent binomial logit models (‘dep.logit’), a probit model (‘probit’), a multinomial logit model (‘multinom’), and the computation of empirical correlations (‘correlation’). The considered covariate is *h* = `Age`; other covariates are considered in Figures A1 to A5. The target disease *j* covers IMID, FMS, onlyIMID, onlyFMS and IMID&FMS and is distinguished by color. Dotted vertical lines represent a difference of zero.

**Figure 3.**
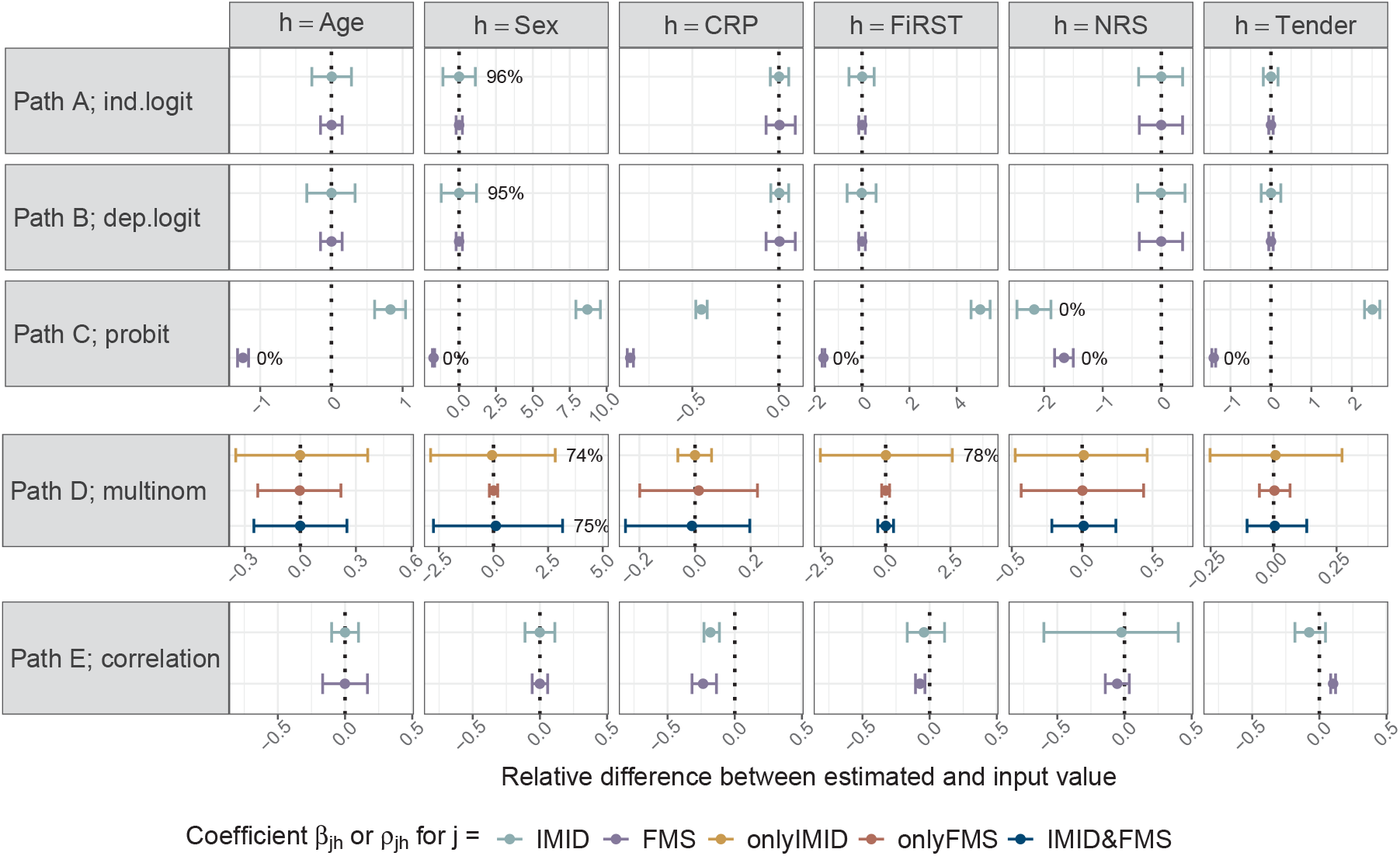
Relative difference between the estimated value of the regression coefficient *β*_*jh*_ or correlation coefficient *ρ*_*jh*_ and the input value. The plots display averages (dots) and 95 % empirical ranges (bars) across 1,000 generated datasets. Percentages indicate how often the signs of estimate and input coincide. Where no percentage is given, it is 100 %. Each row stands for a combination of data generation path and a way to estimate *β*_*jh*_ or *ρ*_*jh*_, respectively (see also Figure 2). Columns represent the six covariates *h*. The target disease *j* covers IMID, FMS, onlyIMID, onlyFMS and IMID&FMS and is distinguished by color. Dotted vertical lines represent a difference of zero.

Regarding the targeted prevalences and associations of single diseases or disease combinations, we compute the difference between the target value and the estimated value for each data generation path and target measure. Figure 4 displays average values across the 1,000 datasets and empirical 95 % ranges.

**Figure 4.**
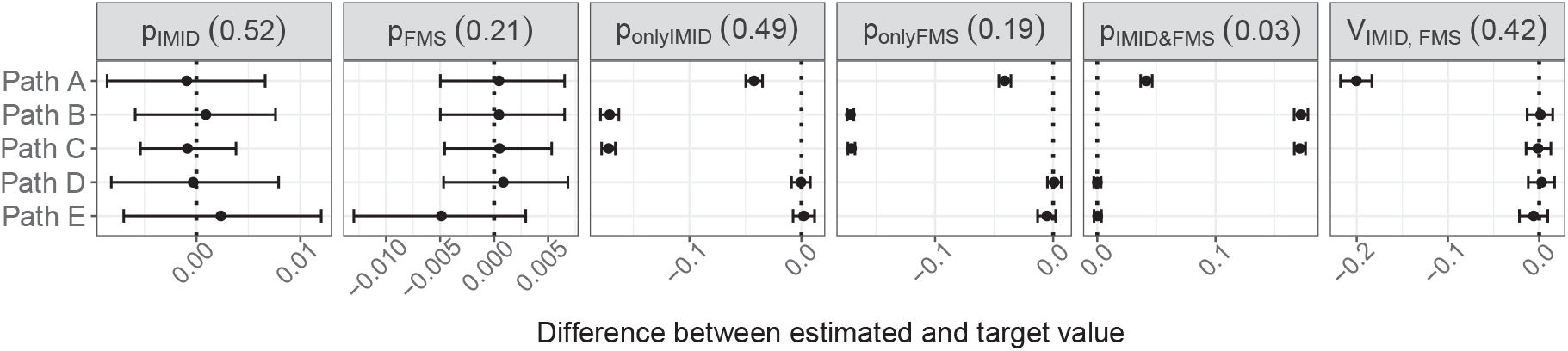
Difference between the estimated values and the target values for all disease (combination) prevalences and Cramer’s V between IMID and FMS. The plots display averages (dots) and 95 % empirical ranges (bars) across 1,000 generated datasets, obtained via Paths A to E. The dotted line represents a difference of zero. In the last plot, the signs of estimate and target coincide agree for every single dataset.

In addition to the graphical representations, we provide tabular overviews of estimated values of regression coefficients and correlation coefficients in Table A4, and empirical disease prevalence and disease association in simulated data in Table A5.

Summarizing the results, we observe that the input information (coefficients and correlations) can be largely recovered from the synthetically generated data if the data-generating process and the estimation method are similar to each other (Figure 3 and main diagonal of Figure 2), while the target properties (prevalences and association between diseases) are achieved to varying degrees (Figure 4).

In particular, it hardly surprises that the regression coefficients generated from Paths A, B and D can be estimated precisely through independent logit models, dependent logit models and the multinomial logit model, where the utilized algorithms for generation and estimation directly correspond to each other. Path C, in contrast, introduces an additional dependence level. It makes use of latent factors, unknown for the estimation procedure, and consequently yields the highest deviations between estimated regression coefficients and input values. It often even fails to maintain the direction of effect. With Path E, the correlation-based generation approach, we can reproduce the input correlations for all covariates with either no or only minor systematic deviation. The sign of the input correlation is maintained for all covariates for all datasets.

The targeted marginal disease prevalence for the single diseases IMID and FMS is obtained with all data generation paths. With respect to the prevalence of disease combinations (that is, onlyIMID, onlyFMS and IMID&FMS), Paths D and E achieve the target and Path A comes comparably close, while Paths B and C show higher deviations.

Regarding the association between IMID and FMS, Path A leads to the largest deviation (in absolute values) for *V*_IMID, FMS_, a consequence of modeling disease occurrence independently, with the only connection between diseases being established via the association with covariates. In contrast, Paths B to E reach the target value to the desired extent. With all paths, the obtained and targeted value of Cramer’s V agree in their direction of association across all synthetic datasets.

## 4 Discussion

We have developed and presented a framework that enables the generation of synthetic data in a medical context involving co-occurring and potentially dependent diseases. Our process consists of four steps: (I) the generation of patient covariates, (II) the calculation of disease predictors based on patient covariates, (III) the calculation of disease probabilities and scores based on predictors, and (IV) the simulation of disease occurrences based on probabilities and scores.

Within each step, various approaches are introduced, which can be combined in a modular fashion. Each of these approaches offer distinct advantages and limitations for specific scenarios. Some of these can be inferred theoretically from the underlying models and were already mentioned in Section 2. The use case from Section 3 further provides insight into a subset of possible applications and practical implications using a concrete data example.

In the following discussion, we focus on the point in the data generation process where the effect of covariates on diseases as well as the dependencies among diseases themselves are modeled and integrated into the data simulation. This corresponds to the transition from Step I to all alternatives within Step II (that is, the model-based Approaches IIa to IId) and the direct transition from Step I to the correlation-based Approach IVe. The Paths A to E in the data simulation in Section 3 were deliberately chosen to represent these five variants. For the remaining parts of the paths, we intentionally kept methodological variation to a minimum in order to better isolate the sources of differences in results.

We describe characteristics of the five Approaches IIa to IId and IVe and compare them in terms of the following aspects:

- **Covariate effects:** How are the intended effects of covariates on (predictors for) disease occurrence included as input into the simulation?
- **Comorbidities:** How does the approach control or induce an association between co-occurring diseases?
- **Causal relations:** What causal structure between diseases is assumed to underlie the data-generating process?
- **Target prevalence:** How is the desired disease prevalence targeted at, and to what extent is it achieved?
- **Use case:** In which context is the approach particularly meaningful?

The following discussion of properties and suitable use cases is also summarized in Table 3.

**Table 3.** Comparison of characteristics of approaches introducing the central dependence structure. **Covariate effects:** How are effects of covariates on disease occurrence included in simulation? **Comorbidities:** How does the approach control or induce an association between diseases? **Causal relations:** What causal structure between diseases is assumed to underlie the data-generating process? Arrows indicate the direction of causation; here exemplary shown for two binary disease variables A and B, covariates ***X*** and latent factor variables ***W***. **Target prevalence:** How is the desired disease prevalence targeted at, and to what extent is it achieved? **Use case:** In which context is the approach particularly meaningful?

| Appr. | Covariate effects | Comorbidities | Causal relations | Target prevalence | Use case |
| --- | --- | --- | --- | --- | --- |
| **IIa** | Directly specified via regression coefficients | Diseases modeled independently; correlation inducible via regression coefficients | No causality between diseases modeled: $\mathbf{X} \rightarrow A$ and $\mathbf{X} \rightarrow B$ | Approximately achieved through intercept (2); precisely achieved through adjustment in IVa | Independent diseases without causal assumption but with co-occurrence due to certain covariates |
| **IIb** | For dependent diseases, specification is blurred due to overlapping model layers | Explicitly modeled via dependent regression models with respective coefficients | Causality through sequentially dependent models: $\mathbf{X} \rightarrow A$ and $\mathbf{X} + A \rightarrow B$ | Approximately achieved through intercept (2); precisely achieved through adjustment in IVa | Sequential dependence of diseases |
| **IIc** | Specification is blurred due to inclusion of latent factors | Specified via inclusion of shared latent factors and respective regression coefficients | Causal effect of shared latent factor on pairs of diseases: $\mathbf{X} \rightarrow A$, $\mathbf{X} \rightarrow B$ and $A \leftarrow \mathbf{W} \rightarrow B$ | Approximately achieved through intercept (2); precisely achieved through adjustment in IVa | Assumption about shared but unknown common cause of disease occurrence |
| **IId** | Directly specified for disease combinations via regression coefficients | Modeled via disease combination categories and odds ratios | Models the distribution of disease combinations; no explicit causality assumption | Approximately achieved through intercept (3); precisely achieved through adjustment in IVb | Knowledge about effect of covariates on occurrence of disease combinations |
| **IVe** | Directly specified via pairwise correlations | Directly specified via pairwise correlations | No causal relation but correlation modeled | Precisely achieved through success probability in marginal binomial distribution | Correlated disease occurrences without causality assumption or knowledge on data-generating process |

In **Approach IIa**, we model disease occurrences as conditionally independent given the covariates. The effect of covariates on each disease predictor can be specified directly by regression coefficients. While we do not explicitly define the relationship between single diseases, a positive or negative correlation can be induced by according choices of regression coefficients; that is, through the values of regression coefficients *β*_*jh*_ and *β*_*lh*_ for different diseases *j* and *l* but identical covariate *h*. The prevalence of each disease can be approximately targeted at by choosing the intercept according to Equation (2), which can be further optimized in Approach IVa. Approach IIa is primarily useful for creating conditionally independent disease occurrences, in particular if we have no suspicion of patient characteristics that promote or exclude a simultaneous occurrence, or of causal relationships between diseases. A limitation of the approach is that target associations between diseases are not directly specified but occur as a result of the impact of covariates. The correlation between diseases is further constrained by the target prevalences of the single diseases.

**Approach IIb** overcomes one limitation of Approach IIa: By generating diseases as a successive sequence, Approach IIb allows to explicitly model disease dependence. In particular, the occurrence of one disease can be modeled to depend not only on patient covariates but also on the presence of one or more other diseases. This comes at the cost of giving up another property, namely the control over effects of covariates on disease occurrence. The final association is blurred by the sequence of model layers, as could be observed in the use case (e. g. Figure 2, Path B). For those diseases that are modeled unconditional on another disease, however, the characteristics resulting from Approach IIa and Approach IIb are equivalent since they use the same underlying model structure. Target prevalences are achieved as in Approach IIa, followed by IVa. Approach IIb is particularly useful if a direct influence from one or several diseases on another one is assumed. The specification of target covariate effects on disease occurrence, however, becomes more complicated.

With **Approach IIc**, we are able to create associations between disease occurrences by including shared latent factors: The occurrence of single diseases is modeled to depend on both individual patient covariates and additional joint latent factors, without the need to define a sequence of dependencies or model a direct causal relation. A key implication of introducing latent factors is that the input regression coefficients are not precisely reproducible from the generated datasets and may even change their sign as observed in the use case example (e. g. Figure 2, Path C). Target prevalences are achieved as in Approaches IIa and IIb, followed by IVa. Approach IIc is particularly useful in cases where diseases are assumed to be dependent, but due to some unknown common cause rather than due to covariate effects as in Approach IIa (or at least not to the same extent). In this context it is important to note that the definition of relationships between diseases in this manner is restricted to the data-generating process; i. e. it does not imply causality between diseases when interpreting the synthetic data, but the unobserved latent factors act as confounders in the underlying structure.

**Approach IId** generates the simultaneous occurrence of diseases by modeling categorical outcomes representing disease combinations, rather than by modeling separate binary outcomes. The effect of covariates on disease combinations is explicitly specified by the regression coefficients and can be precisely reproduced from the generated dataset when applying a multinomial logit model (e. g. Figure 2, Path D). The association between single diseases is induced implicitly rather than specified explicitly. However, the conditional association between two diseases can be specified through odds ratios as explained in Approach IId, Step 1, Option (iii). Approach IId is most appropriate if there is knowledge about the effect of covariates on the occurrence of disease combinations. The prevalence of each disease combination can be approximately targeted at by choosing the intercept according to Equation (3), which can be further optimized in Approach IVb.

**Approach IVe** uses the NORTA algorithm, that means, unlike the previous approaches, it is fully based on specified correlations and marginal distributions instead of using a regression model for simulation. The primary advantage of this approach is its simplicity and the possibility to directly specify the linear relationship between all variables (covariate – disease and disease – disease) via a correlation matrix. Target prevalences are precisely obtained through the indicated success probabilities of the respective marginal binomial distributions of single diseases. If one aims to synthetically generate covariate data through Step Ib (which is NORTA-based as well), Approaches Ib and IVe can be combined to one simulation step to make implementation even easier. Also, Approach IVe can easily be adapted to the scenario of simulating diseases combinations as in Approach IId. As a drawback of Approach IIe, there is no data-generating process modeling the distribution of disease occurrence conditional on covariates or other diseases. While the direction (positive or negative) of the covariate effects on disease occurrence can be influenced through the corresponding correlation, the effect strength is rather induced than specified. Moreover, changing the distribution of covariates while keeping correlations fixed might cause unintended effects on disease occurrence. Overall, Approach IVe is most useful if correlations between diseases and covariates are intended to be strictly controlled but no model is required. In practice, this approach is most applicable if knowledge about correlations is available from real data. In that case, it might be more straightforward to specify a correlation matrix rather than regression coefficients. Still, the approach exhibits limited scalability; as dimensionality increases, selecting a meaningful and valid correlation matrix becomes increasingly difficult.

To summarize, the theoretical considerations and empirical insights show that our modular toolkit provides a wide range of possibilities for generating the desired synthetic data. Thereby, the data generation process is designed to produce synthetic data that serves to validate statistical methods through simulation studies; for example, for the methodological comparison of single-label and multi-label classification which arose from our previous investigation (Schmiegel et al., 2025). Importantly, there are natural limits to fulfilling all wishes for synthetic data (precise specification of covariate effects, disease associations and prevalences) simultaneously. Thus, the user will make a selection of appropriate modules based on the available information and priorities.

## 5 Conclusion

Reliable methods of statistical inference are a fundamental prerequisite for drawing reliable conclusions from medical data. Robust synthetic data, in turn, form the basis for the validation of statistical methods by means of simulation studies. This holds in general, and in particular for the case we consider: the analysis of co-occurring diseases in connection with patient information. We addressed the need for respective data by introducing a four-step framework consisting of several viable approaches for generating synthetic data in case of co-occurring diseases.

Apart from a theoretical classification of the model-based and correlation-based approaches, we tested the framework in practice and applied it to a deliberately chosen selection of five combinations of generation approaches to a specific use case, the co-occurrence of pain-causing diseases. We analyzed the impact of the choice of methodological approaches on covariate effects, association structures and disease prevalence. The approaches differ in their ability to induce specific characteristics regarding covariate effects and disease dependency. We have delineated the strengths and limitations, and recommended use cases for each strategy.

The presented framework for generating synthetic data is intended for the validation of statistical methods, rather than for developing medical therapies or investigating clinical relationships. Thus, though being beneficial, medical realism is not required. The appropriateness of a data generation approach critically depends on the considered use case, in particular, on potential statistical inference techniques and relevant target characteristics of the data. In order to ensure meaningful synthetic data, it remains essential to select modules of the data generation process as well as input and target parameters with careful, informed consideration. Although our work focuses on co-occurring diseases and their association with patient information, the modeling framework is, of course, transferable to any case of simultaneously occurring events and covariate information, also beyond the medical context. The detailed explanations and interpretations offered in this article can serve as general guidelines for researchers regarding synthetic data generation. By offering a structured framework including alternative options and the comparison of multiple approaches, our work provides a valuable contribution to the thorough validation of research methods.

## Data Availability

Due to data privacy protection, we are not allowed to share the data of the German hospital used in this work. However, the code used for synthetic data generation and analysis is available at https://github.com/fuchslab/synthData_diseases.

https://github.com/fuchslab/synthData_diseases

## Declarations

### Computational details

Data generating was done using the statistical software R version 4.4.3 (R Core Team, 2024). The code used for data generation and analysis is available at https://github.com/fuchslab/synthData_diseases.

### Availability of data and materials

Due to data privacy protection, we are not allowed to share the data of the German hospital used in this work.

### Ethics Statement and Consent to Participate

The hospital data used in this work originates from a study for which ethical approval was given by the Ethics Committee of the Westphalia-Lippe Medical Association and the University of Münster (Ethik-Kommission der Ärztekammer Westfalen-Lippe und der Westfälischen Wilhelms-Universität Münster); case number: 2021-426-f-S. All patients agreed on participating in this study.

### Authors’ contributions

H. Marchi and C. Fuchs initiated the project and conceptualized the research goals. H. Marchi conducted literature research, method development and implementation, data analysis and visualization, and drafted the original manuscript. S. Schmiegel and T. Schamberger gave input on structure and methods. All authors edited and critically revised the manuscript.

## Acknowledgements

We would like to thank Martin Rudwaleit and Marvin-Hendrik Röchter for the insightful collaboration in a different research project that introduced us to the medical use case treated here. That project was supported by the Medical Research Start-up Fund of the Medical School OWL, Bielefeld University, and provided patient data. Further, we thank Elmar Spiegel for methodological discussions and valuable feedback.

## A Supplementary Material

### A.1 Derivation of log-odds for disease combinations in multinomial model

In the following, we consider the log-odds of the joint occurrence of diseases, based on information about the exclusive occurrence of single diseases. The derived formulas are employed in Section 2.2.2, Approach IId, Step 1, Option (iii). We focus on the joint consideration of two and three different but potentially related diseases. The derived relationships can also be extended to more than three diseases, and the formulas can be employed as a sub-model in the context of *d >* 3 co-occurring diseases.

To start with, we look at the scenario of two diseases A and B. Let *Y* be a multinomial random variable denoting the disease status of a particular patient which can take the values *A* (meaning that the patient suffers exclusively from disease A), *B* (analogously), *AB* (both diseases occur simultaneously) of *H* (the patient is healthy, the reference category). By *π*_*A*_, *π*_*B*_, *π*_*AB*_ and *π*_*H*_ we denote the probability for *Y* taking the respective value. Additionally, we introduce binary random variables *A* and *B* indicating whether the patient suffers from the respective disease (value one) or not.

The log-odds for *Y* taking the values *A, B* and *AB* are defined as

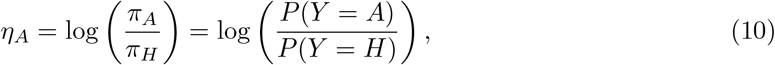

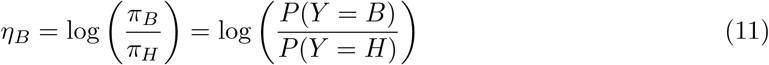

and

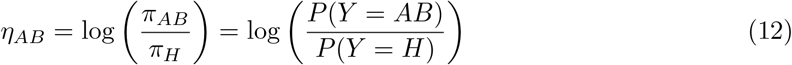

respectively. The odds ratio (OR) for a 2 × 2 contingency table is often used as a measure of association (Sauerbrei and Blettner, 2009). We will make use of it to determine the desired association between two diseases. We first calculate the log-odds ratio for diseases *A* and *B*, using the definition for OR from Agresti (2010), as

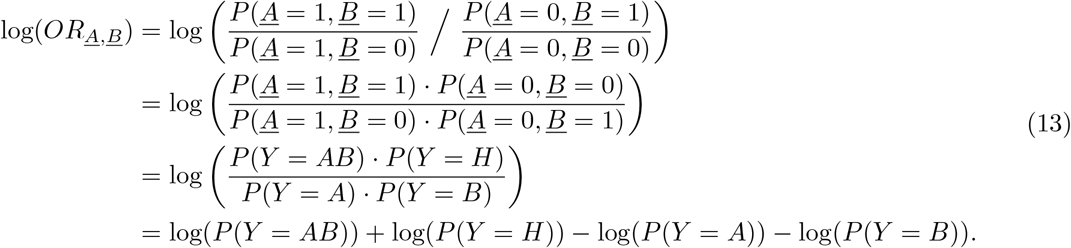

Equations (10), (11) and (12) can be rearranged to the following:

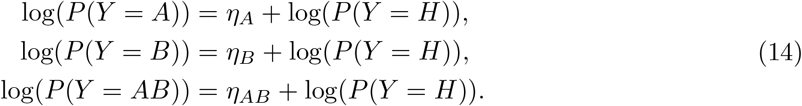

Plugging in the formulas from Equation (14) into Equation (13), we can write log(*OR*_*A,B*_)

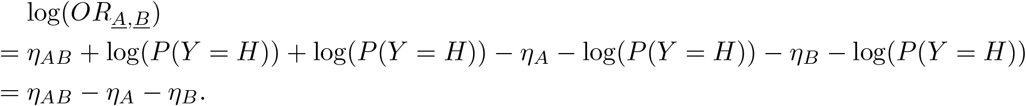

Thus, we can calculate the linear predictor for category *AB* as

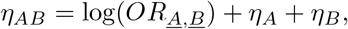

where we set *OR*_*A,B*_ to the desired association between diseases *A* and *B*.

For three diseases A, B and C (and respective definitions of random variables as above), the linear predictor *η*_*ABC*_ for disease combination *ABC* can be calculated as described in the following. The possible disease occurrence combinations are *A, B, C, AB, AC, BC, ABC* and *H*. Additionally to Equations (10)-(12), we define the following:

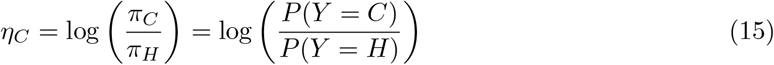

and

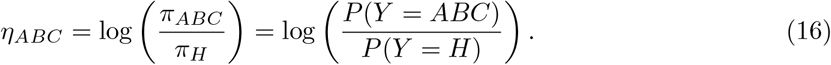

Analogously to Equation (14), we can rearrange *η*_*C*_ and *η*_*ABC*_ accordingly:

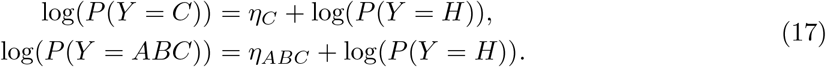

To calculate *η*_*ABC*_ for disease combination *ABC*, we use the log-odds ratio for diseases A, B and C, which can be defined by the ratio of the two conditional log-odds ratios:

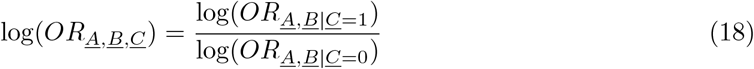

The conditional log-odds ratios can be seen as two different 2 × 2 contingency tables and therefore, similar to Equation (13), calculated as shown here:

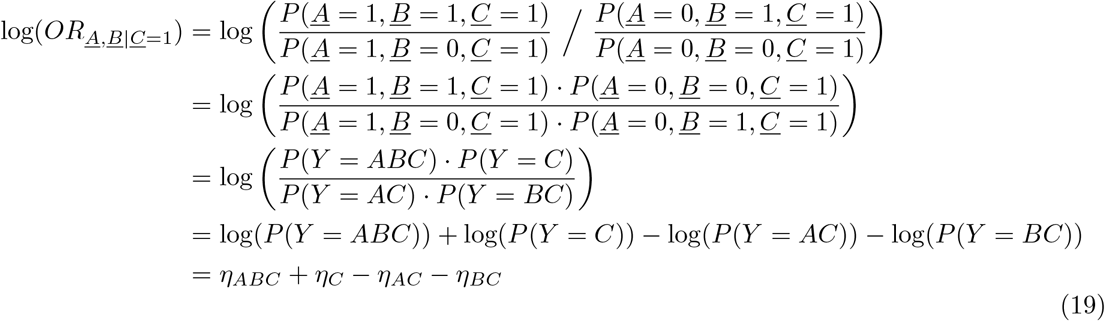

and

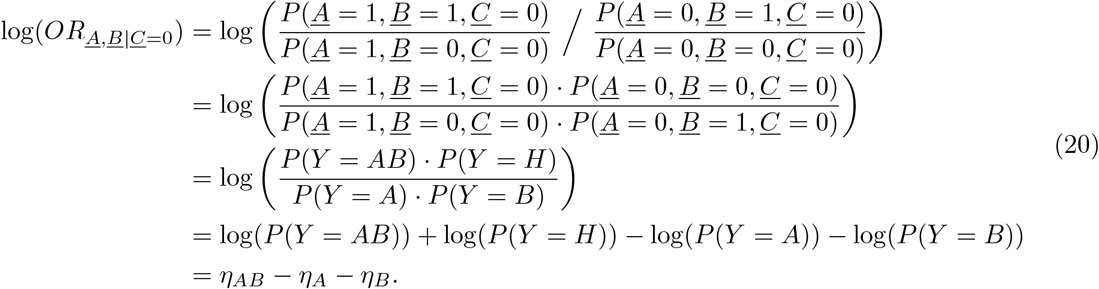

Using Equations (19) and (20) in Equation (18), we get

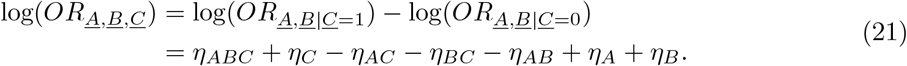

Finally, rearranging Equation (21), the linear predictor for disease combination *ABC* can be defined as:

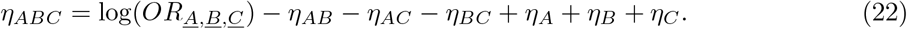

### A.2 Required input specifications for Step II

Table A1 summarizes required input specifications for the model-based Approaches IIa to IId and the correlation-based Approaches Ib and IVe.

**Table A1:** Required input specifications for Approaches IIa to IId and IVe. Here, we assume that Step I has already been completed by using either Approach Ia or Ib. In addition to the listed parameters, one needs to specify prevalences *p*_*j*_ for each individual disease *j* (for Approaches IIa to IIc) or *p*_*r*_ for each disease combination *r* (for Approach IId) to proceed to Steps III and IV.

| Approach | Required parameter |
| --- | --- |
| **IIa** | - Vector of $k$ covariates for patients $i = 1, \dots, n$: $\mathbf{x}_i = (x_{i1}, \dots, x_{ik})'$ (obtained by Step I)<br>- Vector of regression coefficients for each disease $j \in \{1, \dots, d\}$:<br>$\boldsymbol{\beta}_j = (\beta_{j0}, \dots, \beta_{jk})'$ |
| **IIb** | - Vector of $k$ covariates for patients $i = 1, \dots, n$: $\mathbf{x}_i = (x_{i1}, \dots, x_{ik})'$ (obtained by Step I)<br>- Vector of regression coefficients for each disease $j \in \{1, \dots, d\}$:<br>$\boldsymbol{\beta}_j = (\beta_{j0}, \dots, \beta_{jk})'$<br>- Coefficients $\gamma_{jl}$ which describe how the occurrence probability of disease $l$ is influenced by the occurrence of diseases $j \in \{1, \dots, l-1\}$ |
| **IIc** | - Vector of $k$ covariates for patients $i = 1, \dots, n$: $\mathbf{x}_i = (x_{i1}, \dots, x_{ik})'$ (obtained by Step I)<br>- Vector of regression coefficients for each disease $j \in \{1, \dots, d\}$:<br>$\boldsymbol{\beta}_j = (\beta_{j0}, \dots, \beta_{jk})'$<br>- Matrix $\mathbf{B}^*$ defining which diseases $j$ and $l$ share a latent factor for<br>$j, l \in \{1, \dots, d\}$<br>- Set of coefficients $\lambda_{jl} \in \mathbb{R}$ which indicate the influence of the latent factor shared by diseases $j$ and $l$ on disease $j$ |
| **IId** | - Vector of $k$ covariates for patients $i = 1, \dots, n$: $\mathbf{x}_i = (x_{i1}, \dots, x_{ik})'$ (obtained by Step I)<br>- Alternatives:<br>(i) Vector of regression coefficients for each disease combination<br>$r \in \{2, \dots, 2^d\}$: $\boldsymbol{\beta}_r = (\beta_{r0}, \dots, \beta_{rk})'$<br>(ii) Vector of regression coefficients for each disease $j \in \{1, \dots, d\}$:<br>$\boldsymbol{\beta}_j = (\beta_{j0}, \dots, \beta_{jk})'$<br>(iii) Vector of regression coefficients for each single disease $j \in \{1, \dots, d\}$:<br>$\boldsymbol{\beta}_j = (\beta_{j0}, \dots, \beta_{jk})'$; additionally odds ratios $\text{OR}_{ir}$ which quantify for patient $i$ the association between the single diseases contained in combination $r$ |
| **IVe** | - Vector of $k$ covariates for patients $i = 1, \dots, n$: $\mathbf{x}_i = (x_{i1}, \dots, x_{ik})'$ (obtained by Step I)<br>- $(d \times d)$-dimensional matrix $\mathbf{R}_{yy}$ with pairwise correlations between all $d$ diseases<br>- $(d \times k)$-dimensional matrix $\mathbf{R}_{yx}$ with pairwise correlations between all $d$ diseases and $k$ covariates<br>- Marginal cumulative distribution functions $F_{y,j}$ for $j \in \{1, \dots, d\}$<br>- Prevalence $p_j$ for each individual disease $j$ |

### A.3 Parameter values for use case

Tables A2 and A3 list parameters that were derived from real data (with standardized metric covariates) and used as input values for synthetic data generation in the use case in Section 3. These are correlation coefficients among and between covariates and disease occurrences for the correlation-based Approaches Ib (all paths) and IVe (Path E) in Table A2, and regression coefficients for the model-based Approaches IIa to IId (Paths A to D) in Table A3.

**Table A2:** Input correlations for data generation derived from real data. Path E uses the full matrix; for Paths A to D, only rows/columns 3 to 8 are relevant. Correlation was computed as follows: For pairs of variables that are both continuous in the real data, we use Pearson’s correlation coefficient (Pearson, Karl, 1896); for pairs of binary variables, we calculate the tetrachoric correlation (see Section 2.3); for pairs containing one continuous and one binary variable, we calculate the polyserial correlation (Drasgow, 2004, Fox, 2022).

|  | IMID | FMS | Age | Sex | CRP | FiRST | NRS | Tender |
| --- | --- | --- | --- | --- | --- | --- | --- | --- |
| IMID | 1.00 | -0.70 | 0.20 | 0.27 | 0.35 | -0.18 | 0.05 | -0.22 |
| FMS | -0.70 | 1.00 | -0.18 | -0.52 | -0.22 | 0.57 | 0.28 | 0.83 |
| Age | 0.20 | -0.18 | 1.00 | 0.04 | 0.21 | 0.00 | 0.13 | -0.02 |
| Sex | 0.27 | -0.52 | 0.04 | 1.00 | 0.17 | -0.25 | -0.10 | -0.43 |
| CRP | 0.35 | -0.22 | 0.21 | 0.17 | 1.00 | -0.06 | 0.13 | -0.07 |
| FiRST | -0.18 | 0.57 | 0.00 | -0.25 | -0.06 | 1.00 | 0.40 | 0.53 |
| NRS | 0.05 | 0.28 | 0.13 | -0.10 | 0.13 | 0.40 | 1.00 | 0.33 |
| Tender | -0.22 | 0.83 | -0.02 | -0.43 | -0.07 | 0.53 | 0.33 | 1.00 |

**Table A3:**
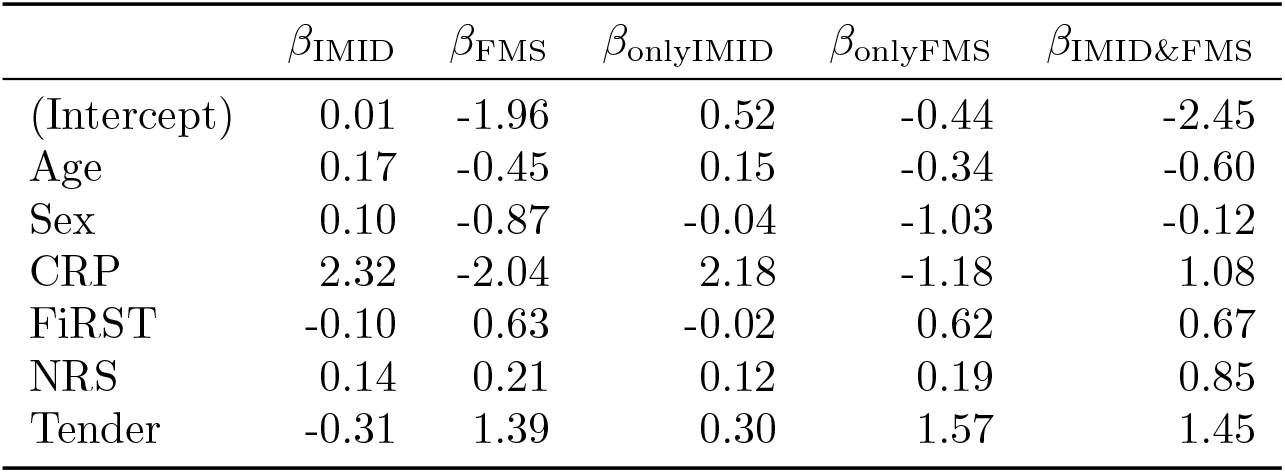
Input regression coefficients for data generation in Step II of Paths A to D, derived from real data as described in Section 3.2.

### A.4 Use case: Additional results

We provide additional results for the use case in Section 3: Just like Figure 2, Figures A1 to A5 display relative differences between the estimated value of the regression coefficient or correlation coefficient and the input value; here for all covariates other than `Age`. Table A4 lists estimated values of the regression coefficient or correlation coefficient, and Table A5 shows empirical disease prevalence and disease association in simulated data.

**Figure A1:**
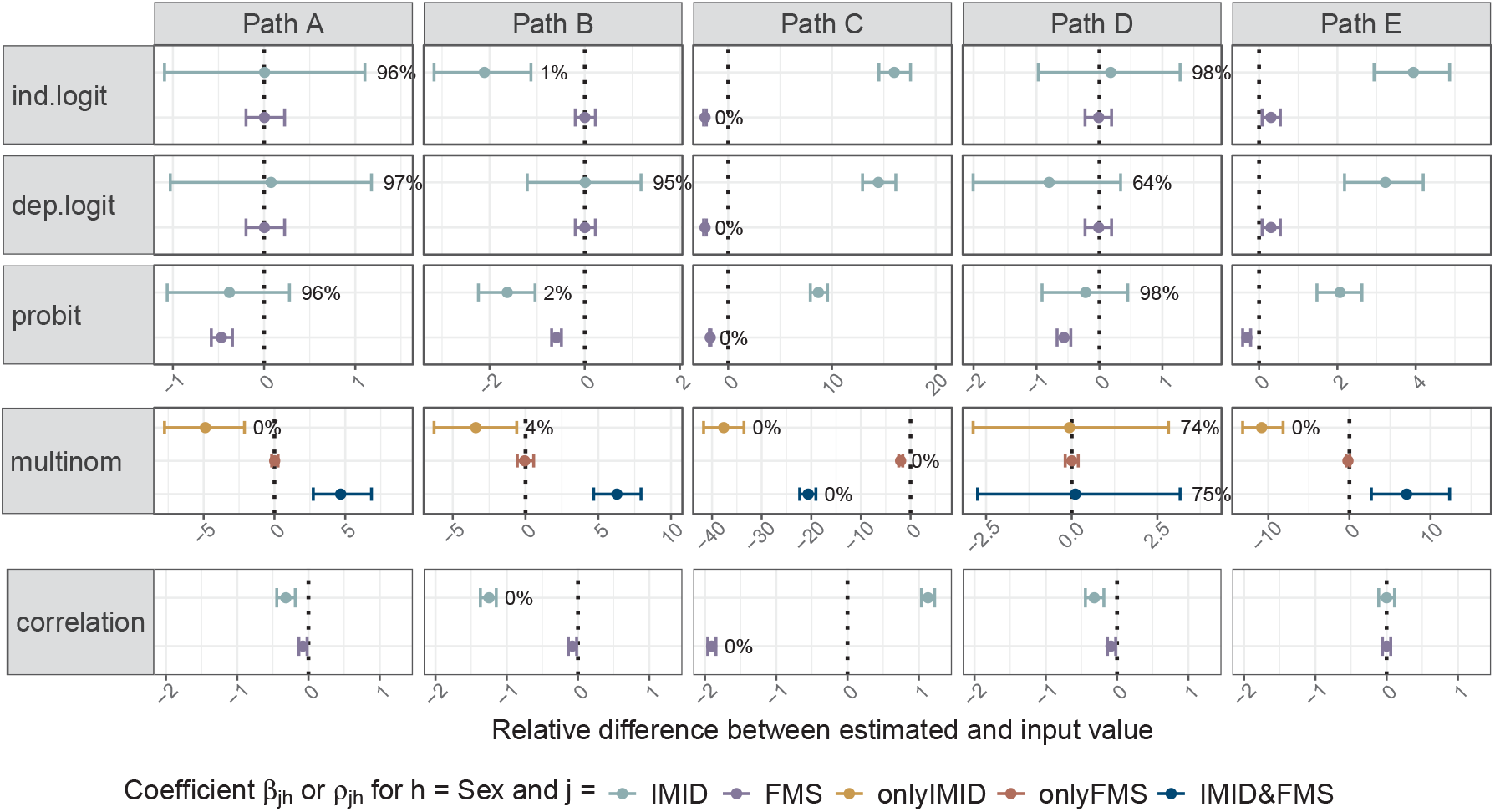
Simulation results as in Figure 2, this time for covariate *h* = `Sex`.

**Figure A2:**
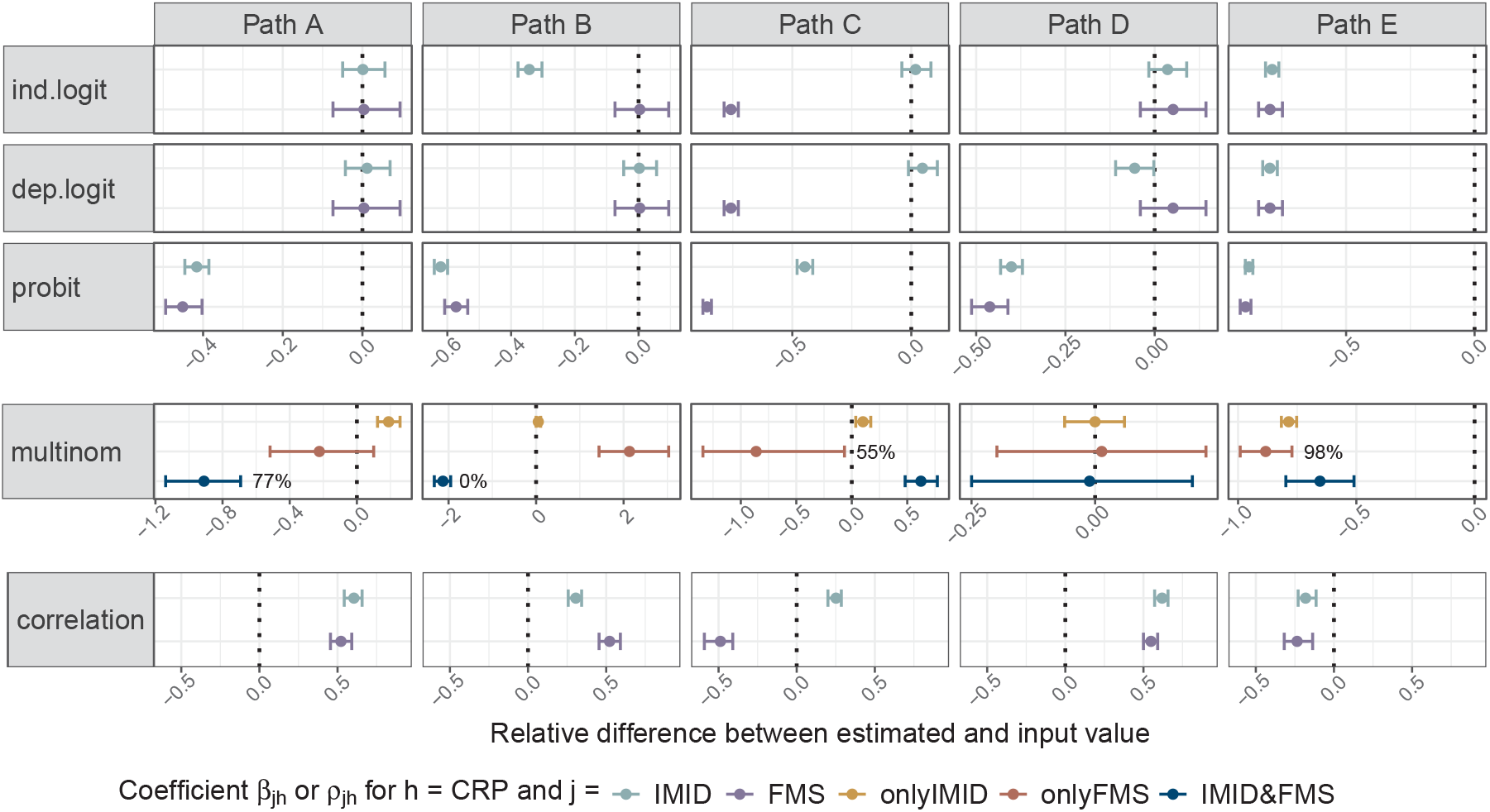
Simulation results as in Figure 2, this time for covariate *h* = `CRP`.

**Figure A3:**
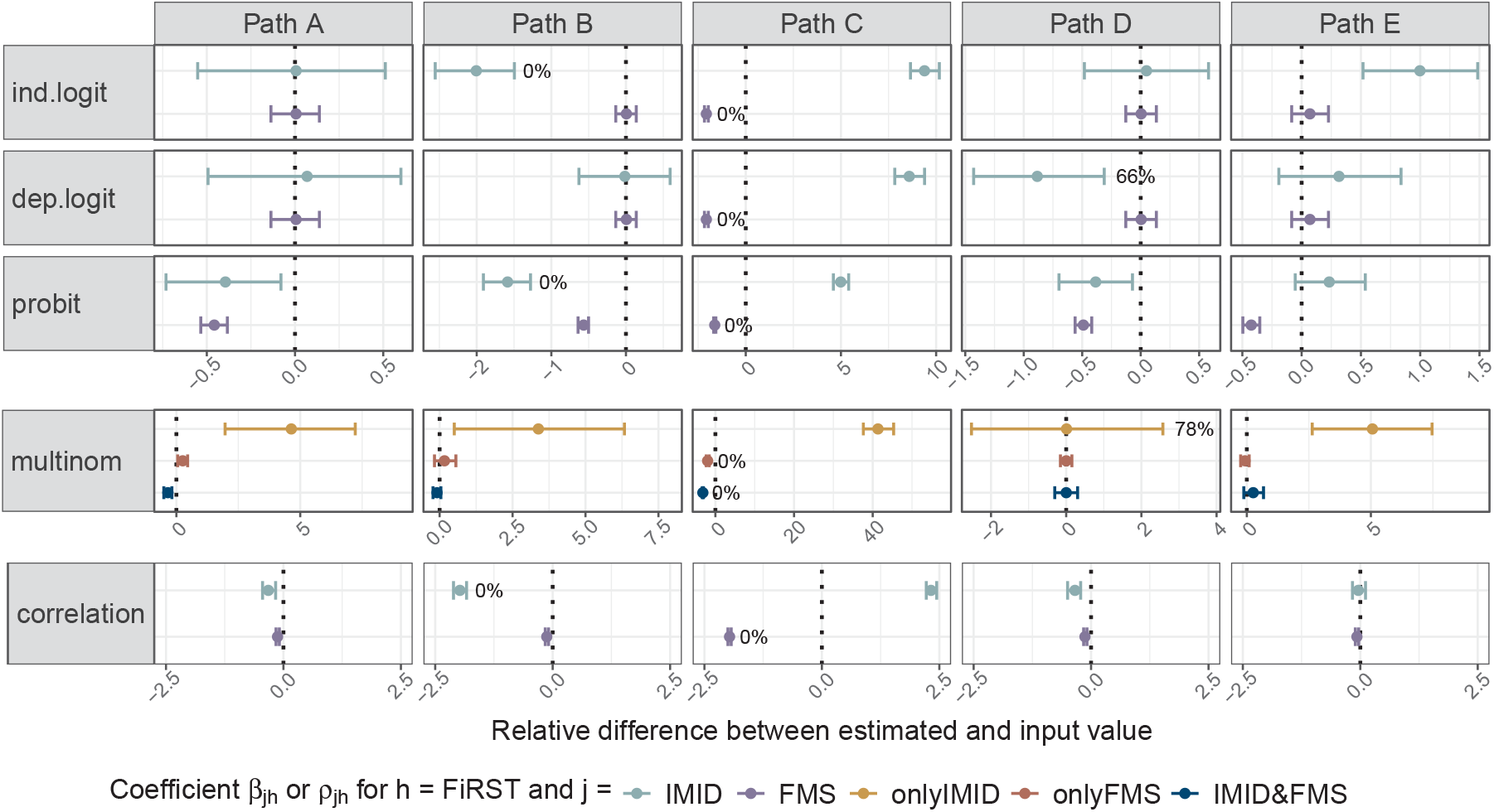
Simulation results as in Figure 2, this time for covariate *h* = `FiRST`.

**Figure A4:**
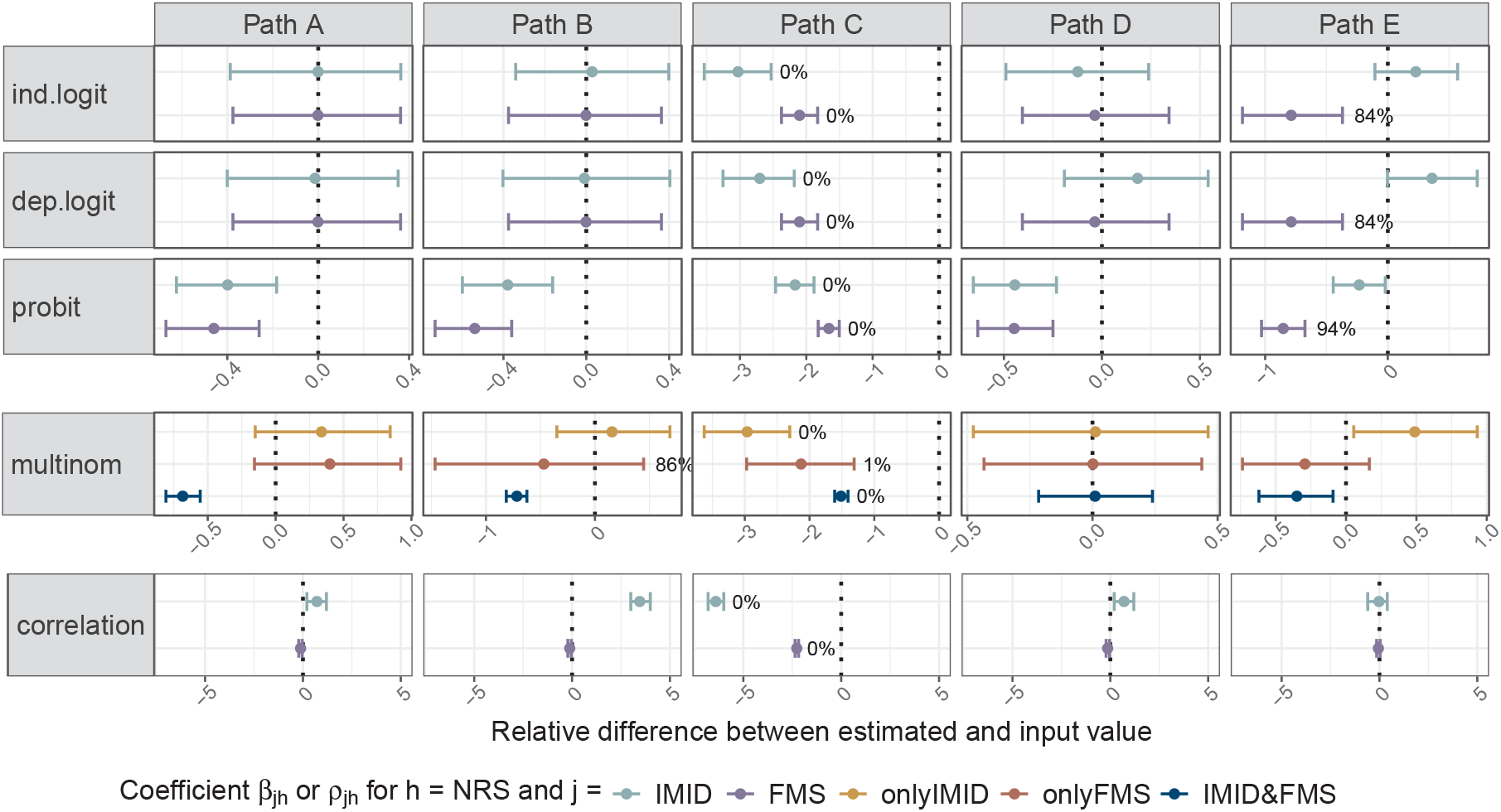
Simulation results as in Figure 2, this time for covariate *h* = `NRS`.

**Figure A5:**
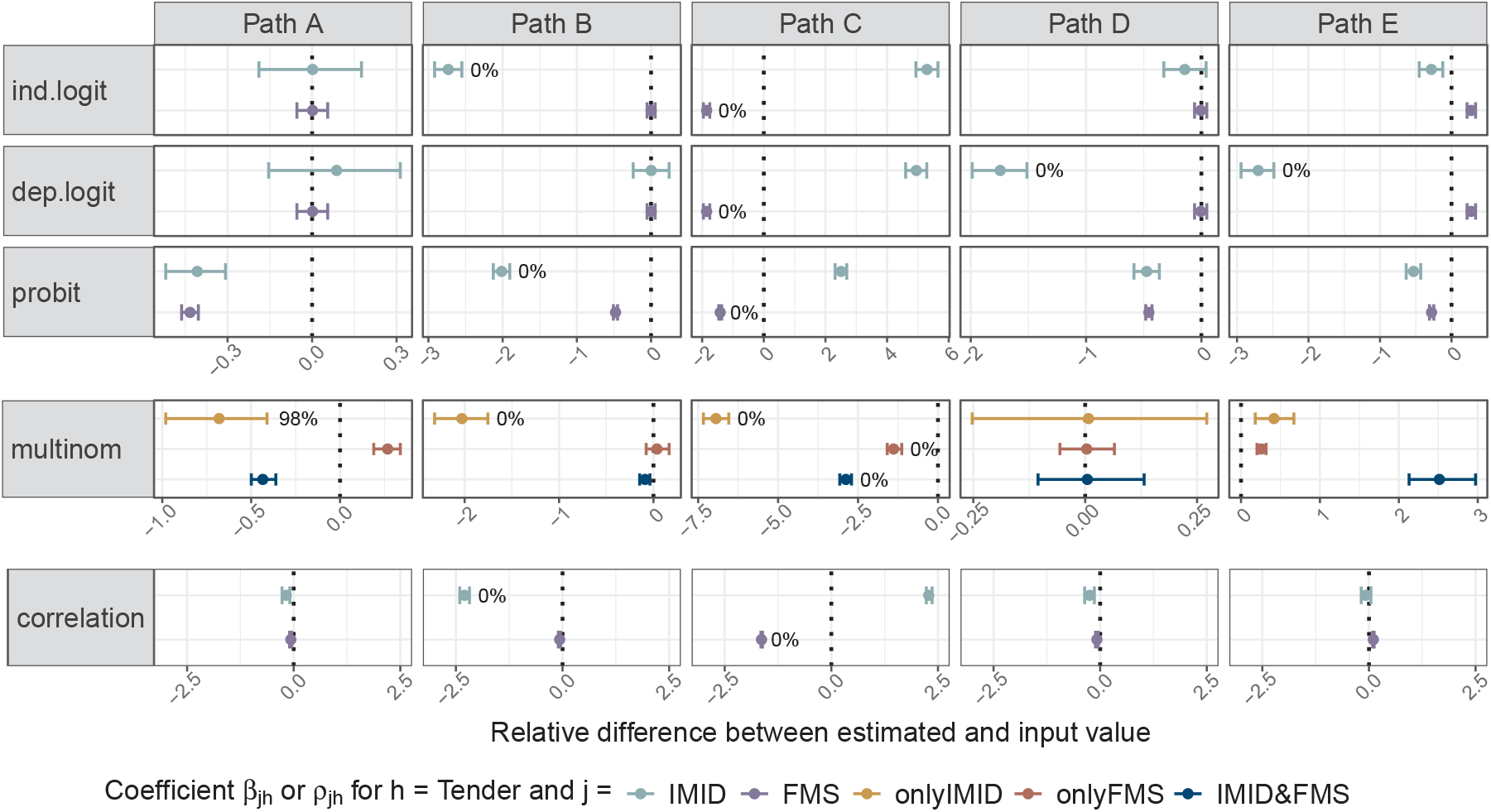
Simulation results as in Figure 2, this time for covariate *h* = `Tender`.

**Table A4:.**
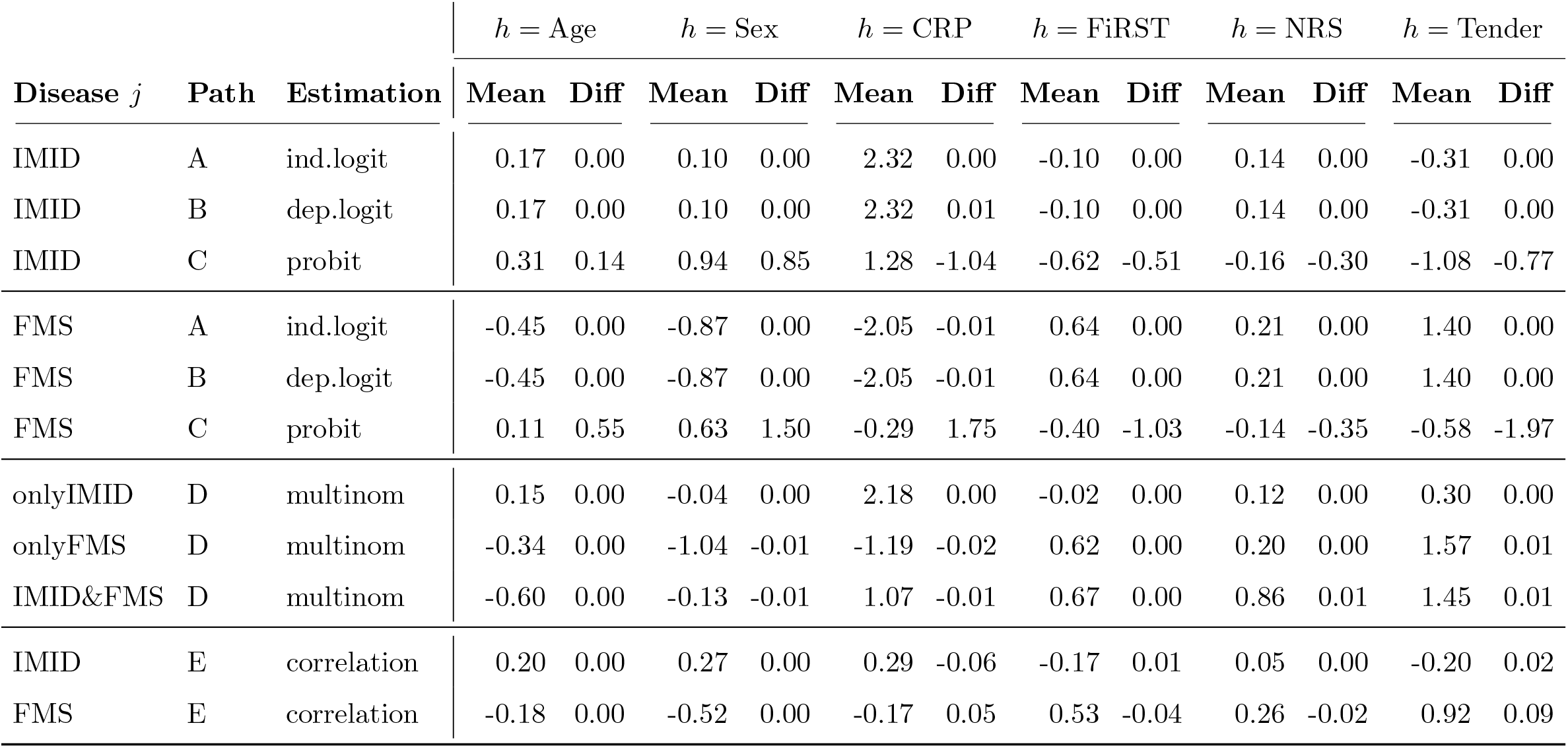
Estimated value of the regression coefficient *β*_*jh*_ or correlation coefficient *ρ*_*jh*_: average values (‘Mean’) and their difference to the input value (‘Diff’) across 1,000 generated datasets. Each row stands for a combination of data generation path and a way to estimate *β*_*jh*_ or *ρ*_*jh*_, respectively (see also Figures 2 and 3). Columns represent the six covariates *h*. The target disease *j* covers IMID, FMS, onlyIMID, onlyFMS and IMID&FMS.

**Table A5:** Empirical disease (combination) prevalence and association in simulated data: average values (‘Mean’) and their difference (‘Diff’) to the target value (‘target’) across 1,000 generated datasets. Each row stands for a data generation path; columns represent the prevalence of diseases or disease combinations (IMID, FMS, onlyIMID, onlyFMS and IMID&FMS) and Cramer’s V between IMID and FMS.

| Path | $p_{\text{IMID}}$ | | $p_{\text{FMS}}$ | | $p_{\text{onlyIMID}}$ | | $p_{\text{onlyFMS}}$ | | $p_{\text{IMID\&FMS}}$ | | $V_{\text{IMID,FMS}}$ | |
| --- | --- | --- | --- | --- | --- | --- | --- | --- | --- | --- | --- | --- |
|  | (target: 0.52) |  | (target: 0.21) |  | (target: 0.49) |  | (target: 0.19) |  | (target: 0.03) |  | (target: 0.42) |  |
|  | Mean | Diff | Mean | Diff | Mean | Diff | Mean | Diff | Mean | Diff | Mean | Diff |
| A | 0.52 | 0 | 0.22 | 0 | 0.45 | -0.04 | 0.15 | -0.04 | 0.07 | 0.04 | 0.22 | -0.20 |
| B | 0.52 | 0 | 0.22 | 0 | 0.32 | -0.17 | 0.02 | -0.17 | 0.20 | 0.17 | 0.42 | 0.00 |
| C | 0.52 | 0 | 0.22 | 0 | 0.32 | -0.17 | 0.02 | -0.17 | 0.20 | 0.17 | 0.42 | 0.00 |
| D | 0.52 | 0 | 0.22 | 0 | 0.49 | 0.00 | 0.19 | 0.00 | 0.03 | 0.00 | 0.42 | 0.00 |
| E | 0.52 | 0 | 0.21 | 0 | 0.49 | 0.00 | 0.18 | -0.01 | 0.03 | 0.00 | 0.41 | -0.01 |

